# Wastewater genomic surveillance tracks the spread of the SARS-CoV-2 Omicron variant across England

**DOI:** 10.1101/2023.02.15.23285942

**Authors:** Franziska S. Brunner, Alexander Payne, Edward Cairns, George Airey, Richard Gregory, Natalie D. Pickwell, Myles Wilson, Matthew Carlile, Nadine Holmes, Verity Hill, Harry Child, Jasmine Tomlinson, Suhel Ahmed, Hubert Denise, William Rowe, Jacob Frazer, Ronny van Aerle, Nicholas Evens, Jonathan Porter, The COVID-19 Genomics UK (COG-UK) Consortium, Kate Templeton, Aaron R. Jeffries, Matt Loose, Steve Paterson

**Affiliations:** Institute of Infection, Veterinary and Ecological Sciences, University of Liverpool CH64 7TE, UK; Deep Seq, Centre for Genetics and Genomics, The University of Nottingham, Queen’s Medical Centre, Nottingham, NG7 2UH, UK; Department of Epidemiology of Microbial Diseases, Yale School of Public Health, New Haven, CT, USA; Biosciences, College of Life and Environmental Sciences, University of Exeter, Exeter EX4 4QD, UK; Environmental Monitoring for Health Protection, UK Health Security Agency, Nobel House, London SW1P 3HX, UK; International Centre of Excellence for Aquatic Animal Health, Cefas, Barrack Road, Weymouth, DT 8UB, UK; Monitoring Laboratories, National Monitoring, Environment Agency EX6 8FD, UK; NHS Lothian, Edinburgh, UK

## Abstract

**Background:** Many countries have moved into a new stage of managing the SARS-CoV-2 pandemic with minimal restrictions and reduced testing in the population, leading to reduced genomic surveillance of virus variants in individuals. Wastewater-based epidemiology (WBE) can provide an alternative means of tracking virus variants in the population but is lacking verifications of its comparability to individual testing data.

**Methods:** We analysed more than 19,000 samples from 524 wastewater sites across England at least twice a week between November 2021 and February 2022, capturing sewage from >70% of the English population. We used amplicon-based sequencing and the phylogeny based de-mixing tool Freyja to estimate SARS-CoV-2 variant frequencies and compared these to the variant dynamics observed in individual testing data from clinical and community settings.

**Findings:** We show that wastewater data can reconstruct the spread of the Omicron variant across England since November 2021 in close detail and aligns closely with epidemiological estimates from individual testing data. We also show the temporal and spatial spread of Omicron within London. Our wastewater data further reliably track the transition between Omicron subvariants BA1 and BA2 in February 2022 at regional and national levels.

**Interpretation:** Our demonstration that WBE can track the fast-paced dynamics of SARS-CoV-2 variant frequencies at a national scale and closely match individual testing data in time shows that WBE can reliably fill the monitoring gap left by reduced individual testing in a more affordable way.

**Funding:** Department of Health and Social Care, UK, Natural Environmental Research Council, UK, COG-UK

**Research in context:** *Evidence before this study:* Genomic monitoring of wastewater for SARS-CoV-2 variants has been introduced in several countries and shown to effectively detect the spread of known variants in multiple studies. However, verification of its alignment with individual testing data at a national scale has so far been reported only for Austria, where sampling covered around 5.4million people. Further and larger scale verifications of the reliability of wastewater-based epidemiology (WBE) are needed to increase confidence in its use for public health monitoring.

*Added value of this study:* We provide evidence that WBE was able to closely track the spread of the emerging SARS-CoV-2 variant Omicron, as well as its sub lineage dynamics, at a regional and national scale across England. Our sampling covered >70% of the English population, equivalent to 39.4 million people. We thereby demonstrate the scalability of our approach to national levels. We also show how WBE is able to track dynamics in different regions of the UK and at a finer scale within London. Its close alignment, in estimated epidemiological timings, with results from intensive individual testing in the same timeframe provides evidence that wastewater-based monitoring can be a reliable alternative when large scale data from individual testing is not available.

*Implications of all the available evidence:* Altogether, evidence is accumulating that WBE is a reliable approach for monitoring SARS-CoV-2 variant dynamics and informing public health measures across spatial scales.

## Introduction

As lockdowns are easing across the world, the threat from SARS-Cov-2, and especially new variants, is still considerable. Therefore reliable, sustainable, and affordable ways to monitor the ongoing evolution of the virus and spread of variants are needed.

Wastewater-based epidemiology (WBE) is a method to monitor pathogens that pose a threat to public health and has been receiving much interest during the SARS-CoV-2 pandemic ^1–3^, exploiting the observation that around 50% of infected individuals shed the virus in faeces ^4,5^.Through sampling of wastewater (sewage) from catchments containing large numbers of individuals, followed by detection of pathogens using molecular methods, WBE can provide regular surveillance of a high percentage of a population ^6^. This provides passive sampling that can encompass individuals who are asymptomatic or who have low engagement with clinical testing programmes ^1,7^. Many local and national WBE programs have been initiated with the prospect of providing insight into the prevalence and diversity of the virus in different communities and to detect the emergence and spread of SARS-CoV-2 variants, for example in Spain ^8^, Switzerland ^9^, India ^10^ and the USA ^11–13^ and have informed local and national public health interventions.

These programs have shown promising results, demonstrating that WBE can detect the occurrence of emerging variants, often even earlier than individual testing ^9,14,15^ and showing differences in the timing of the spread of SARS-CoV-2 variants at a small spatial scale, such as parts of a city ^16,17^. However, WBE verification studies have generally been focused on one or a few cities, with only one published example of SARS-CoV-2 variant monitoring in wastewater at a larger spatial scale across Austria ^18^. The wastewater monitoring program in England had expanded WBE to an unrivalled scale, covering 39.4 million people or 70% of the population by mid-2021 and providing critical insights into uncertainty factors involved with wastewater sampling and their mitigation ^19^. This extensive wastewater sampling coverage, combined with a comprehensive individual testing effort in the UK which can be used for validation, provides an ideal case study to verify the utility of WBE for following the spread of a new variant across a whole country.

The SARS-CoV-2 Omicron variant (B.1.1.529) was first designated as a variant of concern (VOC) by The World Health Organisation on 21st November 2021, following detection in a rapidly growing cluster of South African cases ^20,21^. The Omicron variant is characterised by a large number of mutations, including at sites associated with functional differences on the spike protein, and appears to have arisen from a distinct lineage to that of the previously dominant Delta variant ^22^. Following Omicron’s emergence, it rapidly spread across the globe causing increased Covid-19 case numbers in many countries and was quickly determined to have substantially higher immune evasion than any previous variant, potentially combined with a higher transmissibility ^21^, leading to a strong selective advantage. By mid-January it had already accounted for more than 50% of new infections reported to GISAID and was responsible for more than 90% of new infections detected by individual testing in the UK ^23^. Here we use the arrival and spread of Omicron in England to test the utility of WBE in tracking SARS-CoV-2 variants at a national level. We use genome sequencing of SARS-CoV-2 from >19,000 longitudinal samples and obtained from 524 sites, including sewage treatment works and finer scale sampling from the sewer networks in cities. We use these data to test how quickly WBE was able to detect the arrival of Omicron in England and its ability to then give insight into Omicron’s geographical spread and rise in frequency through time, verifying results against data from individual testing efforts in clinical and non-clinical settings.

## Materials and Methods

### Wastewater Sample Collection and Processing

Wastewater grab samples (1 L per sample) were collected from 233 locations across the sewer network in England and from 291 wastewater treatment plants between the 1^st^ of November 2021 and the 28^th^ of February 2022, as part of the ongoing Environmental Monitoring for Health Protection programme (part of NHS Test & Trace, now the UK Health Security Agency) in England. Samples were transported and subsequently stored at 4 - 6°C until analysis, minimising RNA degradation. Within 24h of collection, all samples were centrifuged (10,000 x g, 20°C, 10 min) in sterile PPCO bottles to remove suspended solids. The supernatant (150 ml) was transferred to 250 ml PPCO bottles containing 60 g of ammonium sulfate (Sigma-Aldrich, Cat. No. A4915). After the ammonium sulfate had dissolved, the samples were incubated at 4 °C for 1 h before further centrifugation (10,000xg, 4 °C, 30 min) and supernatant removal. The pellet was resuspended in 2ml of NucliSens lysis buffer (BioMérieux, Marcy-l’Etoile, France, Cat No. 280134 or 200292). Concentrates were stored at 4°C until nucleic acid extraction. Nucleic acids were extracted from concentrates using NucliSens extraction reagent kit (BioMérieux, Cat. No. 200293) either manually (Farkas et al. 2021) or using the KingFisher 24 Flex system (Thermo Scientific, Waltham, MA, USA) according to the manufacturer instructions ^24^, eluting RNA extracts with a 120 µl volume. Extracts were stored at -80°C until further processing.

### Wastewater Sequencing

Wastewater RNA extracts were purified and sequenced with a standardised EasySeq™ RC-PCR SARS-CoV-2 (Nimagen) V1.0 protocol ^25^. Briefly, samples were cleaned with Mag-Bind® TotalPure NGS beads (Omega Bio-Tek) and then reverse transcribed using LunaScript® RT SuperMix Kit (New England Biolabs) and the EasySeq™ RC-PCR SARS-CoV-2 (novel coronavirus) Whole Genome Sequencing kit v3.0 (NimaGen). Amplicons were pooled and libraries cleaned with Mag-Bind® Total Pure NGS beads (Omega Bio-Tek) or Ampure XP beads (Beckman Coulter) before sequencing on an Illumina NovaSeq™ 6000 or NextSeq 500 platform generating 2×150bp paired end reads. Sample processing and sequencing were performed in 3 different laboratories (Exeter, Liverpool and Nottingham), validated to produce comparable results by sequencing of standardised synthetic samples (Fig. S1). In brief, 47 mixes of 4 different synthetic SARS-CoV-2 RNA Controls (Twist Bioscience) in different proportions were sequenced at each of the sites. We validated reliability between sites by both comparing the frequencies of individual SNPs detected and by comparing the predicted lineage composition from our analysis pipeline from each of the sites with the expected lineage composition based on the known mix of variants.

### Wastewater Bioinformatics

Following sequencing, the wastewater data were demultiplexed and processed using a custom Nextflow v21.04.0 ^26^ pipeline (https://github.com/LooseLab/ww_nf_minimal). Briefly, reads were filtered using fastp v0.20.1 ^27^ and mapped to the SARS-CoV-2 reference (accession: NC_045512.2) using BWA (v0.7.17; http://arxiv.org/abs/1303.3997). Following alignment, primer sequences were trimmed using iVar v1.3.1 ^28^ and a bed file containing the amplicon primer positions. Bam files containing the trimmed alignments then underwent variant (SNP) calling using VarScan v2.4.4 ^29^ and relative SARS-CoV-2 lineage abundance estimation using Freyja (v1.3.1; https://github.com/andersen-lab/Freyja; curated lineage file and UShER global phylogenetic tree downloaded on 2022-07-12). Variant calls were aggregated and subsequent analyses were conducted using custom scripts in R and Python (available on github). The specific SARS-CoV-2 variants (lineages) predicted by Freyja to be present in the samples were summarised into major lineages, i.e. defined variants of concern (VOC) and variants under investigation (VUI) according to the Pango nomenclature (https://cov-lineages.org).

### Sample quality control and method verification

To control for variant detection issues in low quality samples, we excluded wastewater samples from further analyses if coverage depth was ≤ 100x for amplicon 121, which contains 7 of the defining mutations of the Omicron variant, or if less than 100 out of 154 amplicons had at least 20x coverage depth. This reduced our dataset from 19,911 samples to 9,883 samples (Fig S2b).

We further checked these quality thresholds against the SARS-CoV-2 RNA concentration found in the wastewater samples initially by qPCR (Fig.S2c&d).

We used a presence/absence call on characteristic Omicron mutations to validate the results obtained with Freyja for first detection and general detection of Omicron presence in a given area (Fig S3), using the following definitions by Public Health England: The B.1.1.529-BA.1 lineage has 1 synonymous and 16 non-synonymous signature SNPs of which 11 are unique amongst known VOC and VUI, detection confirmed when 11 of 16 signature SNPs and more than 6 of 11 unique SNPs detected and co-occurrence of SNPs on the same amplicon detected, or when more than 11 signature SNPs and more than 6 unique SNPs are present. The B.1.1.529-BA.2 lineage has 4 synonymous and 16 non-synonymous signature SNPs of which 12 are unique amongst known VOC and VUI, detection confirmed when more than 13 of 20 signature SNPs and 6 of 12 unique SNPs detected and co-occurrence on the same amplicon detected, or when more than 13 signature SNPs and more than 6 unique SNPs are present.

We also used Varscan v2.4.4 ^29^ to call SNPs, averaging frequencies across SNPs matching the Omicron profile to validate frequency estimates obtained by Freyja (Fig S4).

### Individual infection sequence metadata

Sequence metadata was obtained from the master COG-UK dataset on 29th March 2022, generated each day using a custom pipeline, datapipe (https://github.com/COG-UK/datapipe). As part of this pipeline, SARS-CoV-2 variants were assigned using Scorpio (https://github.com/cov-lineages/scorpio), and only sequences with sampling dates after the 1st November 2021 and published less than three weeks after the sequencing date were used for Omicron sequences to avoid including incorrectly assigned sequences. The same sample date cut-off as for wastewater samples of 28th February 2021 was used. Geographical locations were also cleaned as part of the analysis pipeline, and details of geographical metadata cleaning can be found here (https://github.com/COG-UK/geography_cleaning). Individual infection data came from two pillars of testing employed in England – pillar 1 data coming from hospital patients and health and care workers, pillar 2 data coming from swab tests in the wider population. We tested the alignment of data from pillar 1 and pillar 2 with wastewater data separately and as pooled data.

### Statistical Analyses

We used R version 4.1.2 ^30^ and RStudio v. 1.4.1717 ^31^ and daily data for all statistical analyses unless stated otherwise. For geographical visualisations of the data, we calculated weekly averages of variant frequencies for each site. For increased visibility on the maps, we then aggregated data by county for both the individual infection data and wastewater data. We plotted maps with complete colouring of the respective counties for better legibility, maps of only the specific areas covered by the sampled wastewater network (covering the main population aggregations) can be found in the Supplement (Fig S5). Shapefiles for mapping were obtained from gadm.org (county delineations) and the maps package in R ^32^. We generated maps of sampling area outlines in Python, using the libraries geopandas v0.11.0, pyplot v3.5.2 and contextily v1.2.0. These outlines are depicted as approximate, not exact outlines due to data sharing restrictions.

We modelled the rise of the Omicron variant in each of the 9 regions of England separately for wastewater and individual infection data by fitting a two-parameter log-logistic function with lower limit 0 and upper limit 1 by least squares estimation using the drm function from the drc package v3.0-1 ^33^.

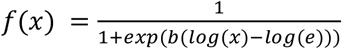

To compare the estimated rates of Omicron spreading in each region and according to different data types, we extracted from each model the date when Omicron was predicted to reach dominance (inflection point, e) and the growth rate during the exponential growth phase which we transformed to a more intuitive growth rate of the function from 5 days before to 5 days past the inflection point as 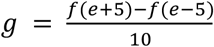

To assess the time match between Omicron frequency estimates in wastewater and individual infection data, we ran Spearman’s correlation tests in the range of time delays of 5 day lags of wastewater data to 3 day lags of individual infection data, using county level daily average frequency data.

## Results

### Detection limits

By comparing synthetic mixes of SARS-CoV-2 variants in each of our sequencing facilities, we could confirm that variant levels as low as 1% were detected as being present in all samples by our sequencing approaches and Freyja based analyses (Fig.S1). Samples with SARS-CoV-2 concentrations above 100,000 genome copies/L consistently yielded sequencing data above our quality criteria, however many samples with lower RNA concentration also yielded sufficient sequencing reads, including many samples where SARS-CoV-2 RNA was not detected by qPCR initially (Fig.S2).

### Detection and frequency estimation of Omicron

We first detected the Omicron variant in wastewater samples from the South East and the East Midlands on the 26th of November 2021, verified with two different approaches using a minimum number of characteristic SNPs and the phylogeny based de-mixing tool Freyja (Fig.S3). The first individual cases from clinical and community testing (referred to as individual testing from here onwards) of Omicron were confirmed by sequencing on the 20th and 22nd of November 2021 in Essex and Greater London, respectively. The following rapid sweep of the variant through England throughout December 2021 (Fig.1) was first evident in wastewater in the London area, where Omicron frequencies reached >25% in the first week of December (Fig.1d) and log-logistic growth models fitted to the regional data indicated that the Omicron variant represented the majority of virus particles in wastewater by the 12th of December (Fig.2a). By contrast, we did not detect Omicron frequencies over 25% in the South West of England until mid December (Fig.1) and dominance of the variant was only predicted from the 21st of December (Fig.2a). We also found a matching decline in the Delta variant across the country, validating our results (Fig.S6). Unfortunately, reduced availability of samples over the Christmas period meant that some areas of the country had no frequency estimates from wastewater samples during the week of the 20^th^ of December. Variant frequency estimates with Freyja were corroborated by estimates based on Varscan, with a maximum discrepancy of 4 days in the estimated date when Omicron reached dominance in any given region (Fig.S4).

**Figure 1:**
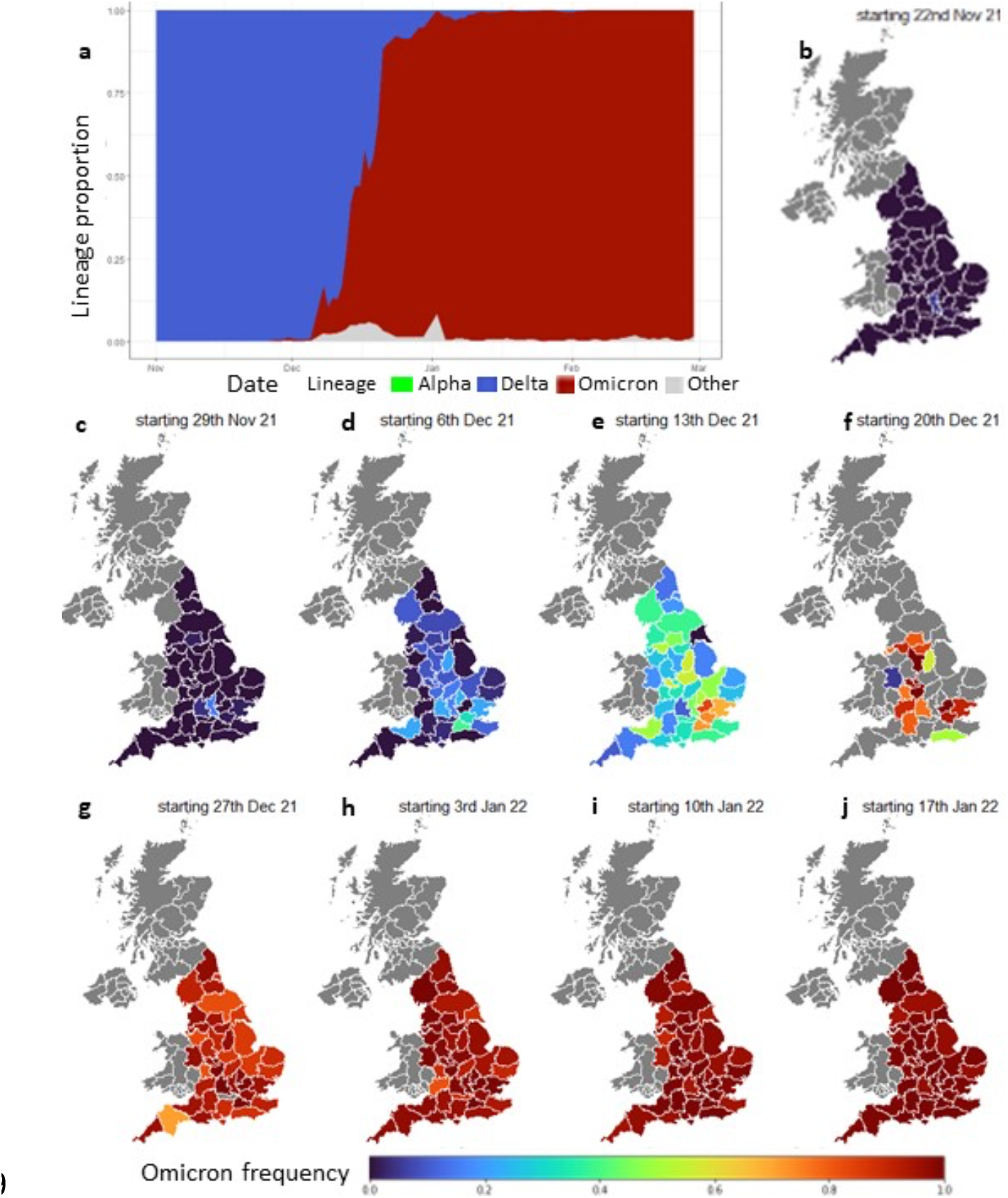
**a** Proportions of major lineages detected in wastewater across England between November 2021 and February 2022. Proportions are shown as 3-day rolling averages weighted by sample numbers. ‘Other’ indicates data remaining unassigned by Freyja to either Alpha, Delta or Omicron. **b-j** Wastewater detection of Omicron variant frequency, averaged by county and calendar week, between late November 2021 and mid-January 2022. Grey areas indicate missing data, due to low sample quality or lack of sampling.

**Figure 2:**
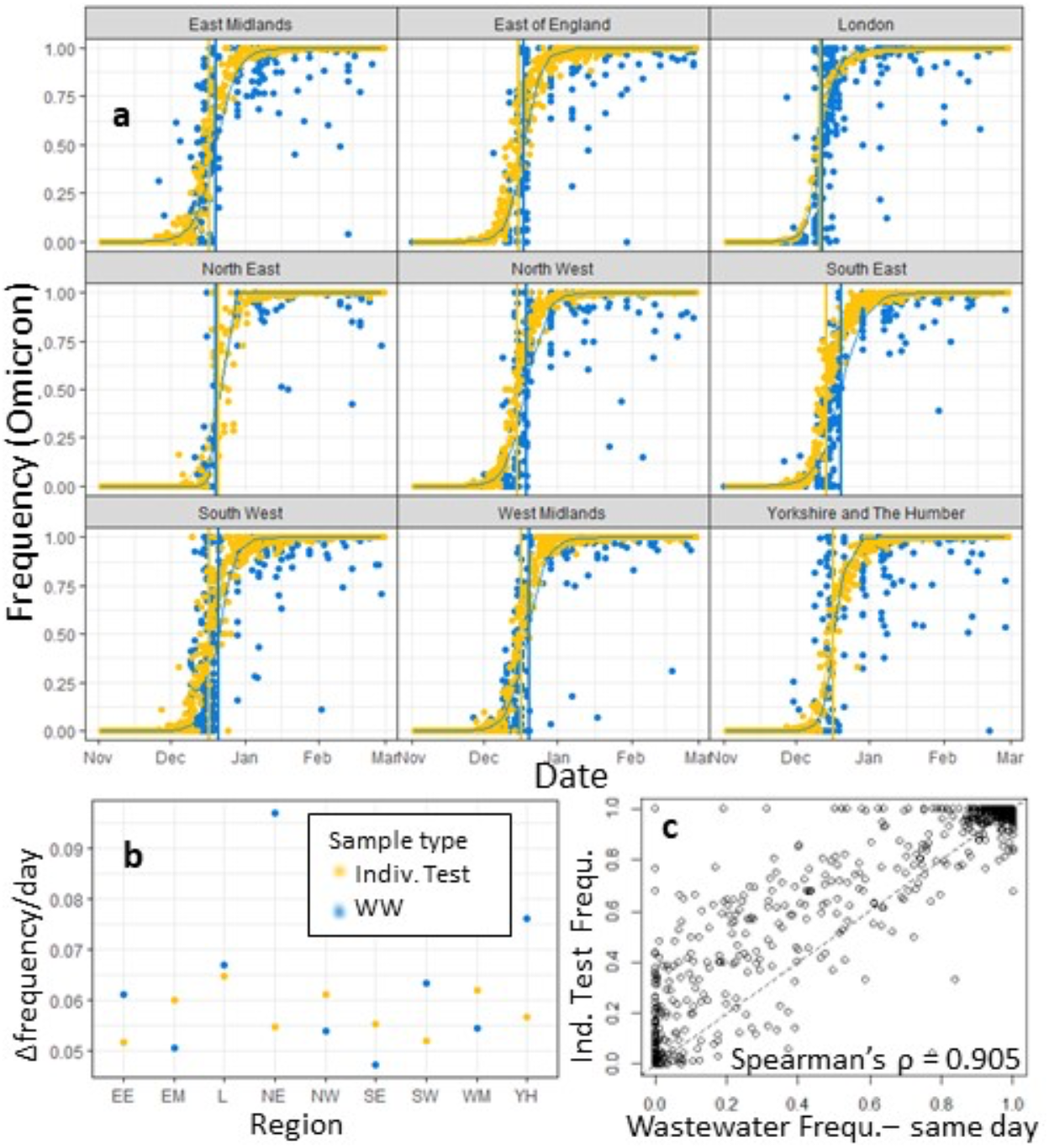
**a** Log-logistic growth models of the Omicron frequencies in 9 English regions. Vertical lines indicate the inflection points of the models which estimate the timepoints when Omicron frequencies reach 50% in wastewater (blue) and individual testing (yellow) samples. **b** growth rates of the log-logistic growth models in each region in wastewater (blue) and individual testing (yellow) samples, calculated as predicted frequency change between 5 days before and 5 days after inflection point. **c** Spearman’s correlation tests show strongest correlation between county level daily Omicron frequency in individual testing and wastewater with no time lag between frequency estimates.

A closer look at the London area also revealed finer scale differences in the spread of Omicron (Fig.3). The first notable levels of Omicron appeared in the East of the city in the week of the 29th of November, but subsequently the central parts of the city saw a quicker rise in Omicron prevalence to over 25% in the week of the 6th of December and also earlier full dominance (>50% frequency) of the variant in the following week.

**Figure 3:**
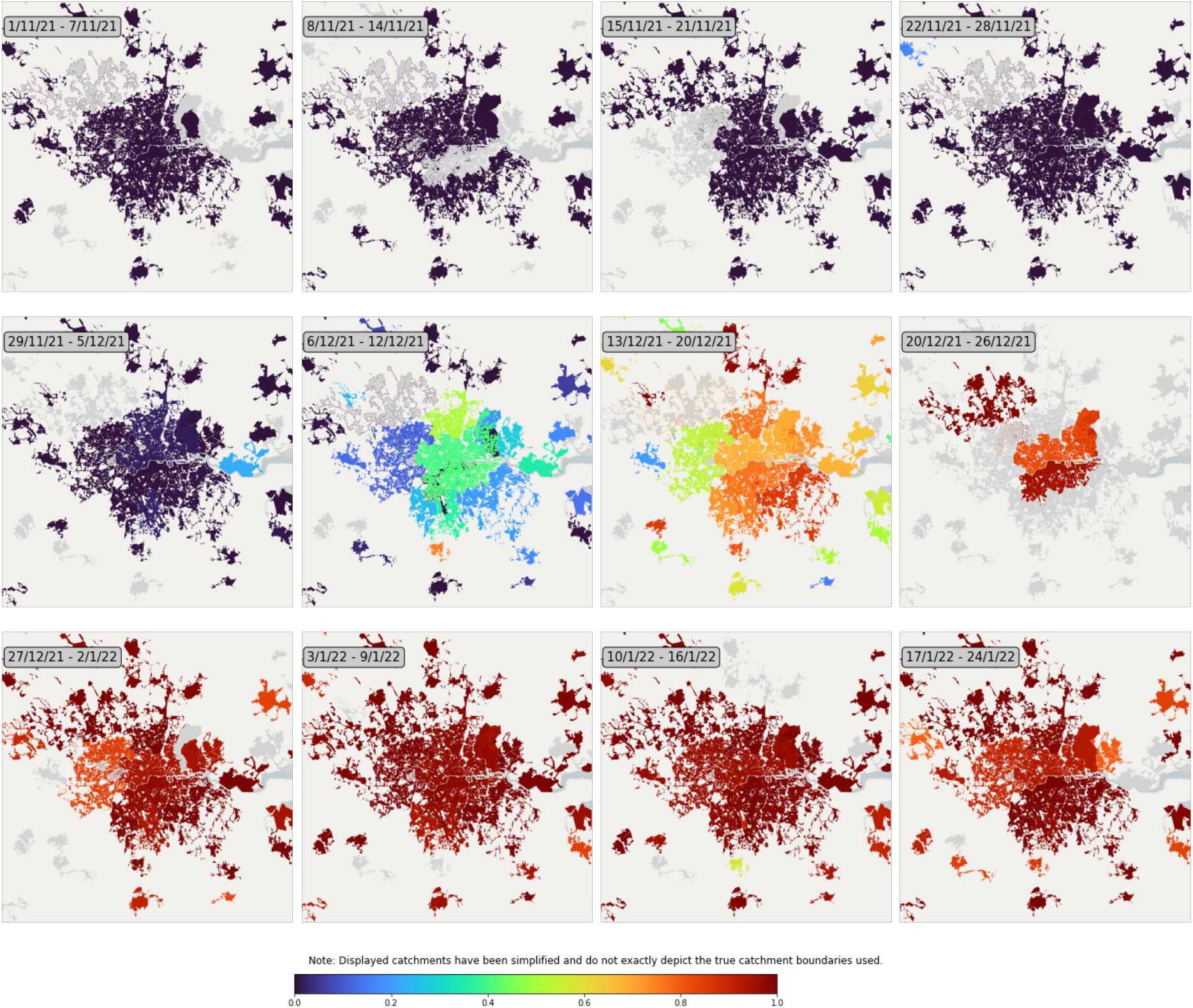
Weekly Omicron frequencies within London. Depicted are approximate outlines of the wastewater catchment area captured by sampling. Grey areas indicate missing data, due to low sample quality or lack of sampling.

### Matching wastewater base results and individual testing results

Individual testing data from clinical and community PCR testing closely matched our wastewater results, with the Omicron variant becoming the majority of detections in the London area by mid-December and the rest of England following suit about a week later (Fig.2). When modelling the logistic growth of Omicron frequencies at the region level, the time point estimates for Omicron becoming the dominant variant in wastewater varied between a 1 day lead on individual testing estimates (North East) to a 6 day lag (South East) (Fig.2a). Frequency growth rate estimates for Omicron were higher in wastewater data in five of nine regions, with rates of 0.052-0.065 frequency change/day in individual testing and 0.047-0.097 frequency change/day in wastewater (Fig.2b). Growth rate estimates were highest for wastewater data from the North East, however this was also the data subset with lowest sample numbers around the transition phase in December, making the rate estimate less reliable. Daily Omicron frequencies aggregated at county level correlated strongly between individual testing and wastewater data across a range of investigated timing lags (Table S1), with the strongest correlation observed when no lag was assumed between wastewater and individual testing data (Spearman’s ***ρ*** = 0.905, Fig.2c). Examination of the different types of individual testing data from Pillar 1 data (collected from patients and staff in hospitals) and Pillar 2 data (collected in the community outside hospitals) revealed a good alignment with wastewater estimates in both data types (Fig.S7). Pillar 1 data yielded slightly earlier estimates of Omicron dominance, with a predicted lead time of 6 days or more on wastewater data in six out of nine regions, whereas Pillar 2 data was predicted to lead on wastewater data by 6 days in only one region (Fig.S7a). Combining data for all of England, we found the strongest correlation for a 2 day lag of wastewater based frequencies behind Pillar 1 data (Spearman’s ***ρ*** = 0.871, Fig.S7b). We found greater variability between regions in the estimates for Pillar 1 data, which is likely related to much lower sample numbers and thereby increased stochasticity.

### Sublineage patterns

We further tested the sensitivity of our lineage detection in wastewater by reconstructing the shift from subvariant BA1 to subvariant BA2 of the Omicron lineage and found a clear shifting pattern from BA1 to BA2 in wastewater samples for each of the nine regions through February (Fig.4).

**Figure 4:**
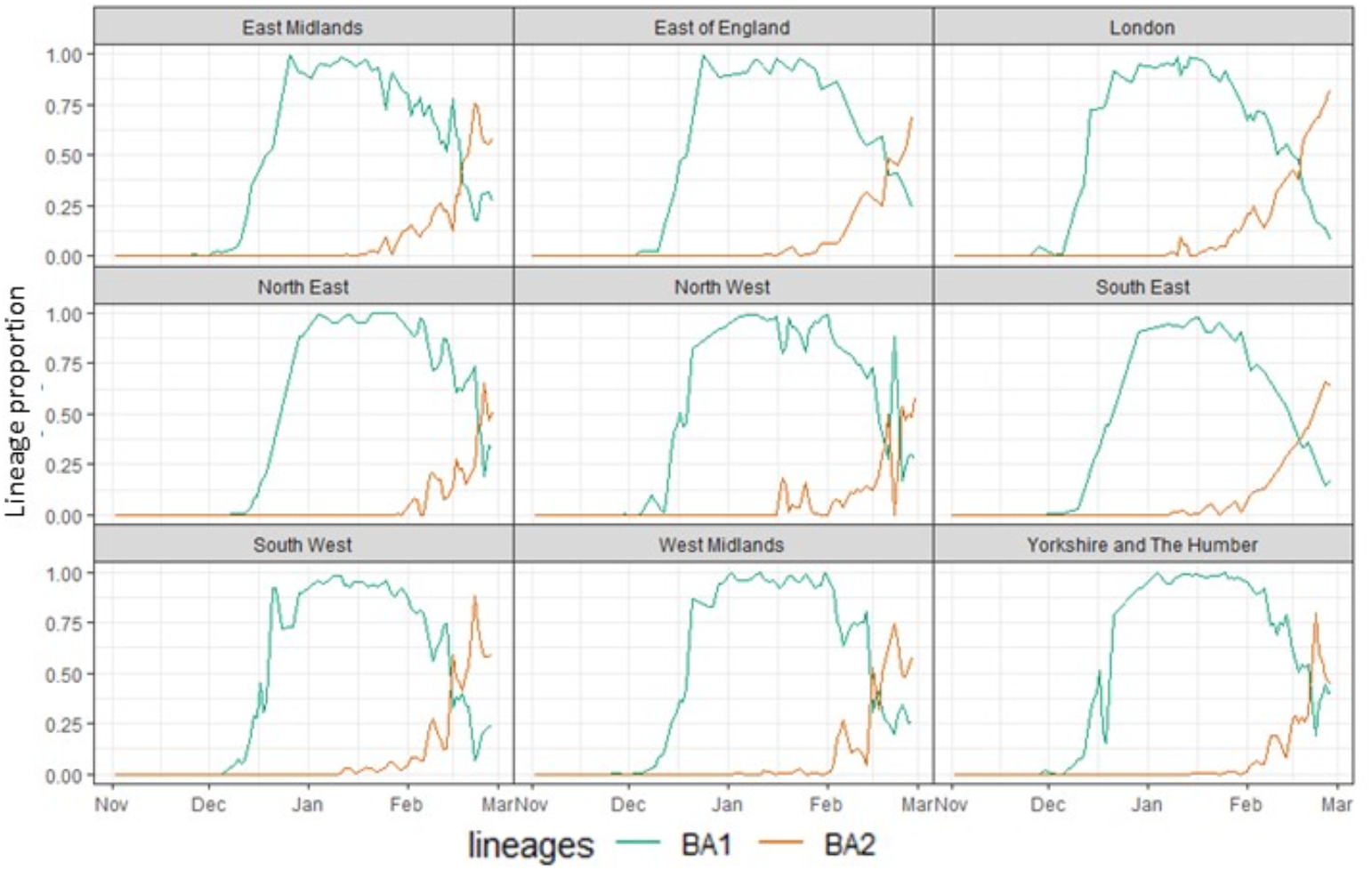
Frequencies of the subvariants BA1 and BA2 of the Omicron variant between November 2021 and February 2022, shown as 3 day rolling averages of frequencies out of all detected variants in wastewater samples.

## Discussion

We show that tracking of the SARS-CoV-2 Omicron variant by WBE closely mirrors that seen by mass community surveillance (i.e., the sequencing of PCR positive individuals) and mapped the rise of Omicron through time, with the earliest rise seen in London followed by the rest of England. Estimated Omicron frequencies from the two approaches are highly correlated, with no overall lag time between WBE and community surveillance by sample date, despite small differences in estimated growth rates of Omicron and transition timings from Delta to Omicron when looking at different regions within England. If we consider only samples taken in a hospital setting, we see an even earlier rise of the Omicron variant, however this comparison is weakened by the much more sparse data from the hospital environment compared to wider community testing. Furthermore, we show that even sublineage dynamics such as the transition from Omicron subvariant BA1 to BA2 during February 2022 ^34^ can readily be detected with our WBE approach.

The ability to detect the presence of an emerging variant – regardless of frequency – may be of particular utility for local public health teams. WBE has the advantage that it is able to monitor infections in areas exhibiting low engagement with community testing programmes, or where not all PCR-positive infections are genome sequenced or from asymptomatic infections ^7^. On the other hand, WBE has disadvantages in a lack of individually identifiable information, which limits the potential for public health teams to investigate where cases live or work or whether they are recent travellers to an area. Here we find that initial detections differ between individual testing and WBE. For example, the first instances of detecting the Omicron variant in November 2021 in wastewater were in the East Midlands and South East whereas the first individual infections with the variant were confirmed in Essex and Greater London. This is perhaps unsurprising given that detecting the first cases of a new variant in an area, either by individual testing or WBE, is inherently stochastic. Using both techniques would therefore maximise the likelihood of capturing these early events. During late 2021, the UK also tested travellers entering the country, with the aim to detect and prevent the spread of imported variants. However, in March 2022 all travel restrictions into the UK were removed and individual testing in the wider community (Pillar 2 approach in England) reduced to minimal levels, in an approach similar to that of many other countries. Our data indicate that WBE could be used to detect the initial spread of variants from around 1% prevalence. The use of WBE could therefore represent a viable alternative for genomic surveillance with minimal bias, low costs and demonstrated sensitivity.

Our demonstration of a close alignment in nation-wide WBE and individual testing data also shows that some inherent differences between these two types of samples may be less relevant than previously thought. Wastewater samples capture mostly faecal shedding of virus particles and a meta-analysis has shown that only an average of 43% of Covid-19 patients shed detectable levels of faecal SARS-CoV-2 RNA and that the duration of particle shedding in faeces exceeds that in respiratory samples by weeks ^5^. If this impacted the proportion between SARS-CoV-2 variants, we would expect to see different variant dynamics between individual sampling and wastewater, especially in the transition phase when a new variant spreads across a country. However, our data show a close alignment in variant dynamics nonetheless, suggesting that no major bias in shedding exists between variants at least and that WBE provides a robust measure of variant proliferation at local and national scales.

The comparative logistic requirements of community sampling and WBE merit discussion. The data presented here represent around 1k-2.3k samples per week for wastewater sampling and around 30k-48k samples per week for community sampling, which come from RNA left over from PCR testing. In the context of the UK testing programme at the time, 1-3.5m PCR tests were conducted each week, of which around 10-32% were positive. Testing at this national scale is a substantial effort that is likely impossible to sustain indefinitely as SARS-CoV-2 becomes endemic and has already been abolished in many countries. Sequencing from only clinical samples, e.g. hospitalised patients, is an alternative, but this may bias sampling against younger age-groups; who are least likely to be hospitalised but among whom transmission may be highest. Furthermore, reduced individual surveillance leads to later detection of newly emerging or imported variants in a country. Wastewater sampling has a significant set up cost, in developing agreements with water companies, installation of autosamplers and logistics to transport samples to laboratories and then in overcoming technical challenges of extracting SARS-Cov-2 RNA from wastewater at scale. On the other hand, it can provide high coverage of a population using fewer resources than, and without the bias associated with, community testing. Around 70% of England was covered by the WBE analysis presented here involving sampling over 200 sewage network site and close to 300 sewage treatment plants. Optimisation of sampling site distribution could potentially provide similar high coverage of the population while reducing the number of sites sampled. As shown in samples from the London sewer network, it also provides insights into the emergence or rise of new variants and how this varies across a city. This means that wastewater monitoring of SARS-CoV-2 evolution is a feasible and cost-effective option for keeping track of emerging local hotspots of the disease and the spread of new variants, even when large-scale individual testing is wound down. The wastewater samples used, and the general approach of targeted pathogen sequencing can also be used for ongoing surveillance of other pathogens to help protect public health, as evidenced already for polio and monkeypox ^35,36^.

Overall, we have demonstrated the usefulness and reliability of WBE for tracking an emerging variant from its first detection to its spread across a whole country. This method can therefore be considered a reliable tool for monitoring large scale genetic dynamics in wide-spread diseases.

## Data Availability

Wastewater sequencing data are publicly available on the European Nucleotide Archive under Study ID PRJEB55313. The clinical case data used in this study are visualised at https://www.cogconsortium.uk/tools-analysis/public-data-analysis-2/. A filtered, privacy conserving version of the lineage-LTLA-week dataset is publicly available online (https://covid19.sanger.ac.uk/downloads) and gives access to almost all used data, despite a small number of cells having been suppressed to conserve patient privacy.

https://www.ebi.ac.uk/ena/browser/view/PRJEB55313

## Acknowledgements

We thank B. Jones, R. Crompton, T. Foster and N. Cadu for assistance with laboratory work. We thank partners across the Environmental Monitoring for Health Protection programme (EMHP) in England for the coordination and collection of wastewater samples. This includes the UK Health Security Agency (UKHSA), water companies in England, the Environment Agency (EA), Centre for Environment, Fisheries and Aquaculture Science (Cefas) and the Department for Environment, Food and Rural Affairs (Defra).

Funding was provided by NERC (NE/V003860/1) and DHSC UK (2020_097). This report is independent research funded by the Department of Health and Social Care. COG-UK is supported by funding from the Medical Research Council (MRC) part of UK Research & Innovation (UKRI), the National Institute of Health Research (NIHR) [grant code: MC_PC_19027], and Genome Research Limited, operating as the Wellcome Sanger Institute. The authors acknowledge use of data generated through the COVID-19 Genomics Programme funded by the Department of Health and Social Care. The views expressed are those of the authors and not necessarily those of the Department of Health and Social Care or UKHSA.

## CRediT Author contributions

**Franziska S Brunner** - conceptualisation, data curation, formal analysis, investigation, methodology, project administration, validation, visualisation, writing – original draft, writing – review & editing

**Alexander Payne** - conceptualisation, data curation, investigation, methodology, resources, software, validation, writing – review & editing

**Edward Cairns** - investigation, methodology, project administration

**George Airey** - investigation, methodology, project administration

**Richard Gregory** - project administration, resources, software

**Natalie D Pickwell** - investigation, methodology, project administration

**Nadine Holmes** – investigation, methodology

**Myles Wilson** - investigation, methodology

**Matthew Carlile** - investigation, methodology

**Verity Hill** - investigation, methodology, software, writing – review & editing

**Harry Child** - investigation, methodology, project administration

**Jasmine Tomlinson** - investigation, methodology

**Suhel Ahmed** - investigation, methodology

**Hubert Denise** - investigation, methodology, writing – review & editing

**William Rowe** - investigation, methodology

**Jacob Frazer** - investigation, methodology

**Ronny van Aerle** - investigation, methodology, writing – review & editing

**Nicholas Evens** - investigation, methodology, supervision, writing – review & editing

**Jonathan Porter** - investigation, methodology, supervision, writing – review & editing

**The COVID-19 Genomics UK (COG-UK) Consortium** - funding acquisition, investigation, methodology, project administration, resources, software

**Kate Templeton** - conceptualisation, funding acquisition, supervision

**Aaron R. Jeffries** - conceptualisation, funding acquisition, investigation, methodology, supervision, writing – review & editing

**Matt Loose** - conceptualisation, funding acquisition, investigation, methodology, supervision, writing – review & editing

**Steve Paterson** - conceptualisation, funding acquisition, investigation, methodology, supervision, writing – review & editing

## Data accessibility statement

Wastewater sequencing data will be made publicly available on the European Nucleotide Archive under Study ID PRJEB55313. The clinical case data used in this study are visualised at https://www.cogconsortium.uk/tools-analysis/public-data-analysis-2/. A filtered, privacy conserving version of the lineage-LTLA-week dataset is publicly available online (https://covid19.sanger.ac.uk/downloads) and gives access to almost all used data, despite a small number of cells having been suppressed to conserve patient privacy.

## Ethics Statement

Use of surplus nucleic acid derived from routine diagnostics and associated patient data was approved through the COG-UK consortium by the Public Health England Research Ethics and Governance Group (R&D NR0195).

