## Supplemental material for "Wastewater genomic surveillance tracks the spread of the SARS-CoV-2 Omicron variant across England"

[8]https://www.cogconsortium.uk, Full list of consortium names and affiliations are in the appendix

[9]NHS Lothian, Edinburgh, UK

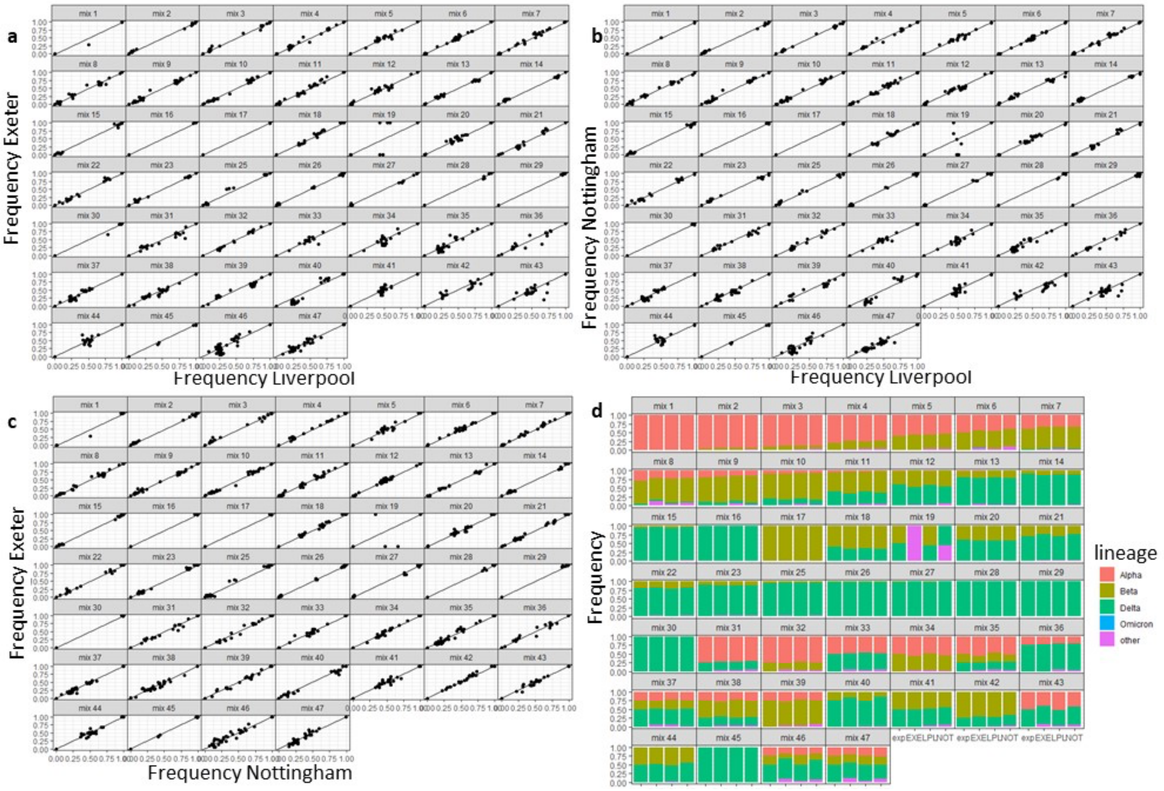


Figure S1: Validation of results comparability from three laboratories using synthetic samples. **a-c** pairwise comparisons of results from two sequencing sites across 47 mixes of 4 synthetic SARS-CoV-2 strains. Each dot shows the observed SNP frequencies in sequencing results from each site. Lines show expected correlation of 1 **d** comparison of expected frequencies of lineages (exp) and lineage frequencies predicted by Freyja based on sequencing results from Exeter (EXE), Liverpool (LPL) and Nottingham (NOT) across 47 mixes of 4 synthetic SARS-CoV-2 strains.

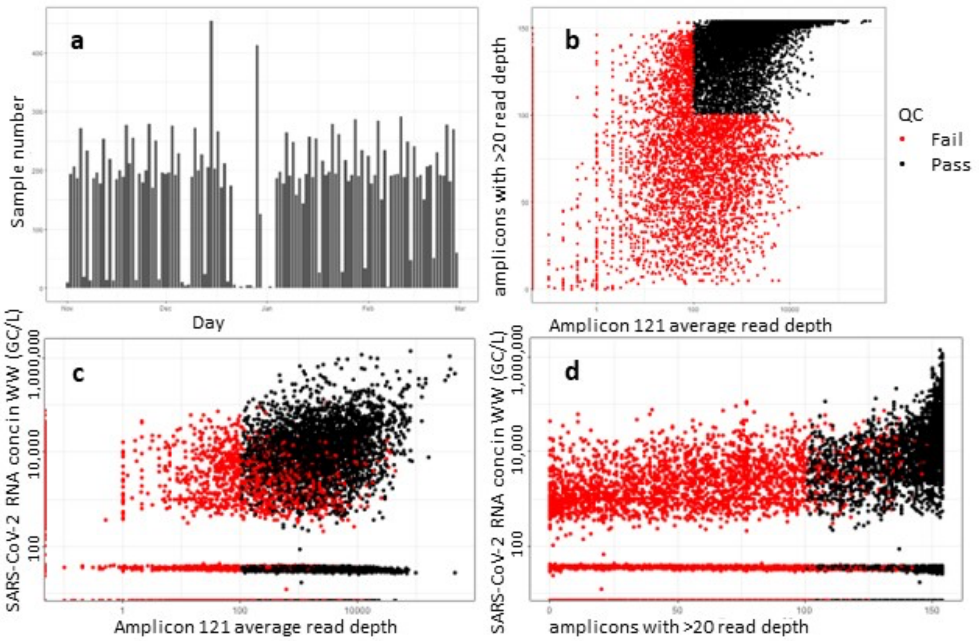


Figure S2: Sample numbers and quality control. **a** wastewater sample numbers per day, **b** cut-offs for our 2 sample quality criteria - greater than 100x average read depth on amplicon 121

and more than 100 (out of 154) amplicons with read depth over 20, **c** SARS-CoV-2 RNA concentration in wastewater versus average read depth on amplicon 121, **d** SARS-CoV-2 RNA concentration in wastewater versus number of amplicons with read depth over 20

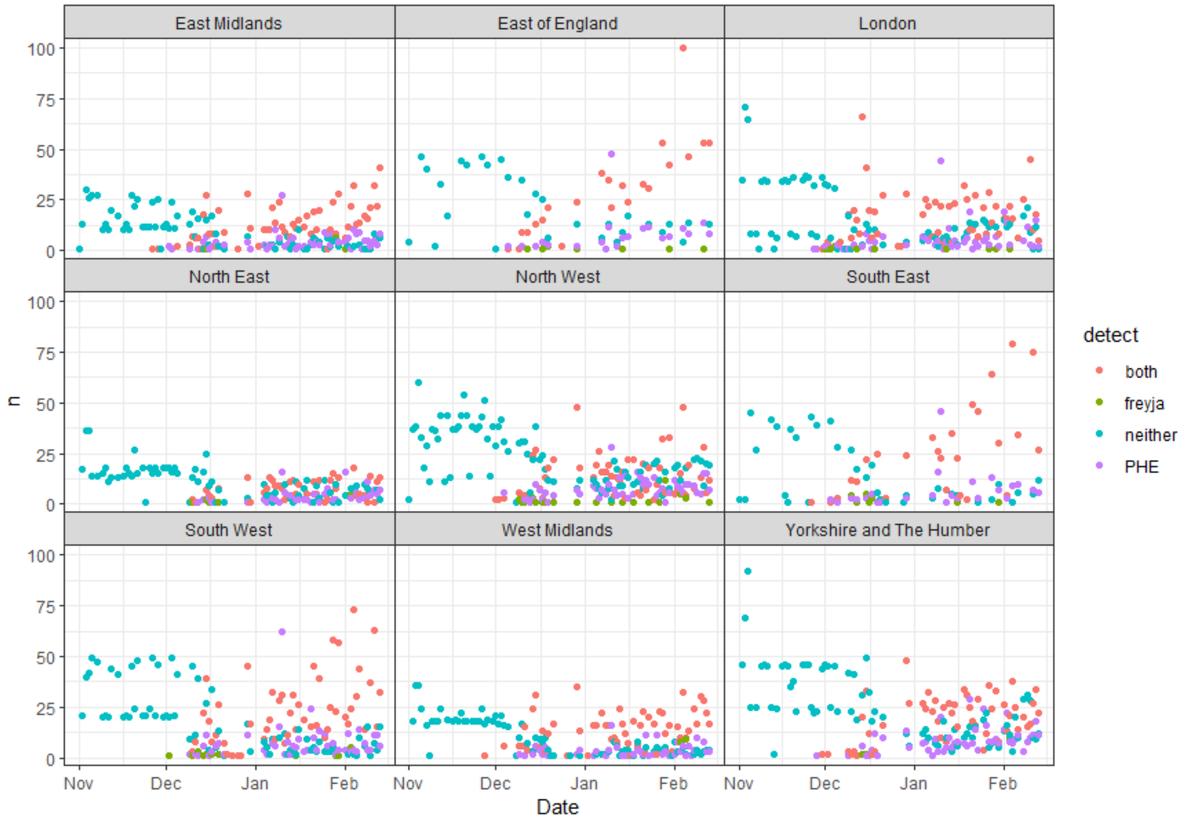


Figure S3: Detection of Omicron by different methods. Detections are shown as counted detections across all wastewater sites sampled in a region on a given date. PHE= detection based on Public Health England definition, freyja= detection by the phylogeny based de-mixing tool Freyja

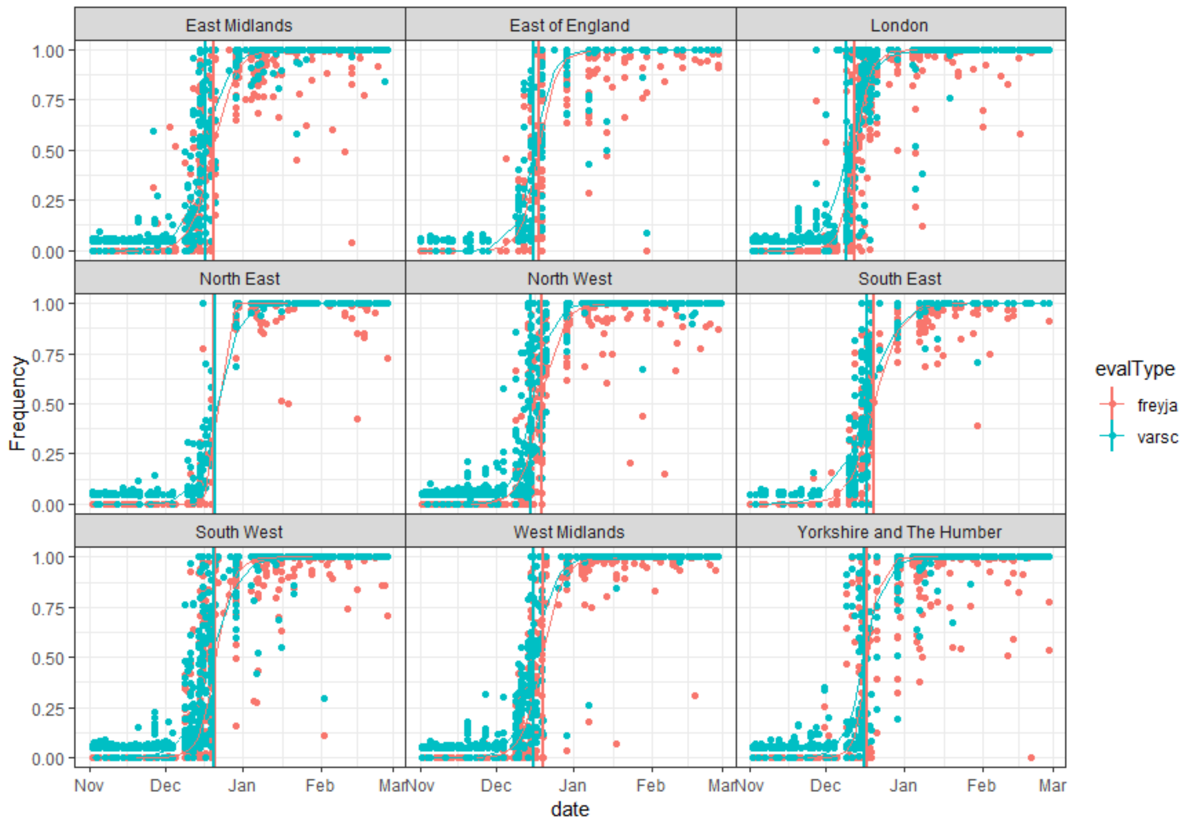


Figure S4: Omicron frequency estimation with individual SNP frequencies (using Varscan) and with the phylogeny based de-mixing tool Freyja. Log-logistic models for each frequency estimation method and region are plotted and vertical lines show the inflection points of these models, which is the estimated timepoint when the Omicron variant reached 50% in the given region.

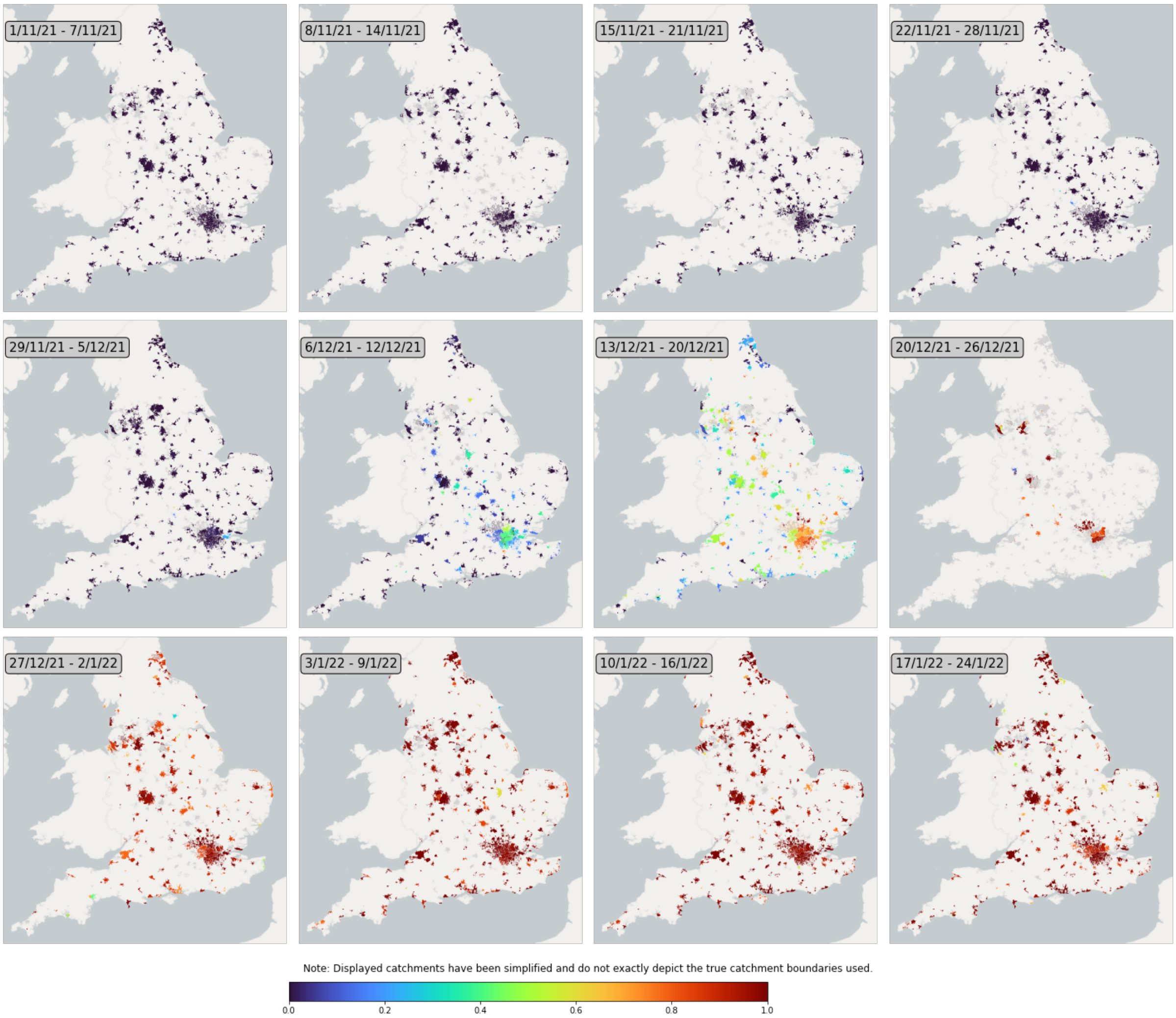


Figure S5: Catchment projection of Omicron detection in wastewater per week. Depicted are approximate outlines of the wastewater catchment areas captured by sampling.

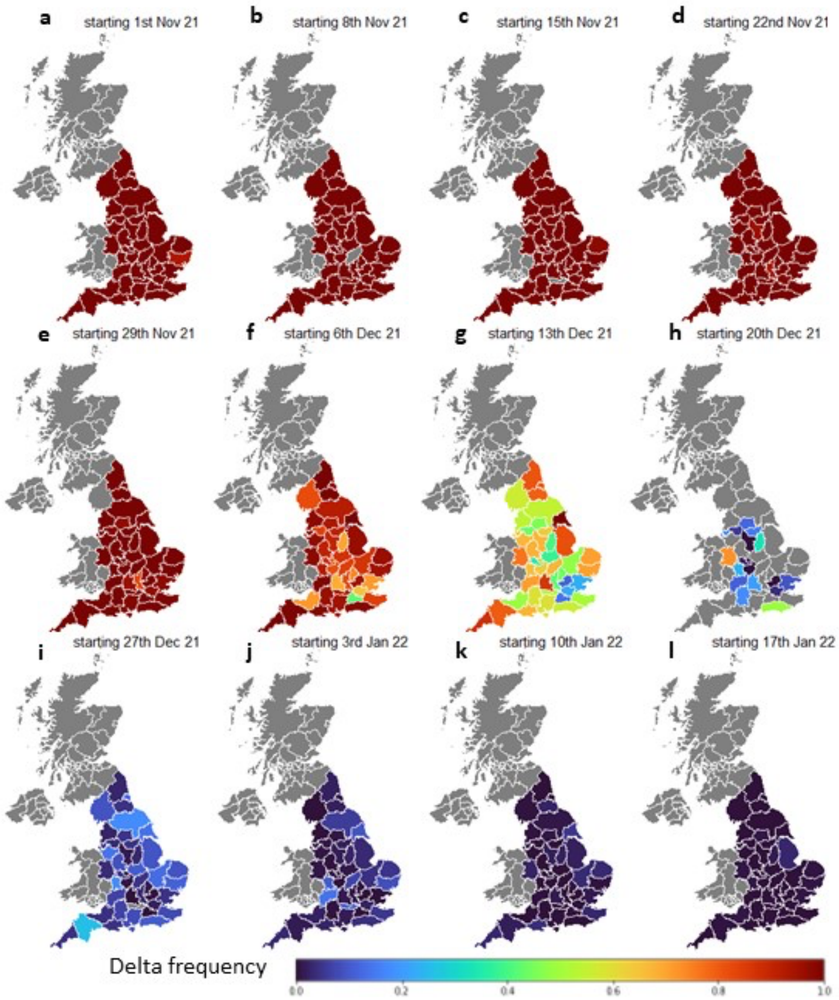


Figure S6: Wastewater detection of Delta variant frequency, averaged by county and calendar week. Grey areas indicate missing data, due to low sample quality or lack of sampling.

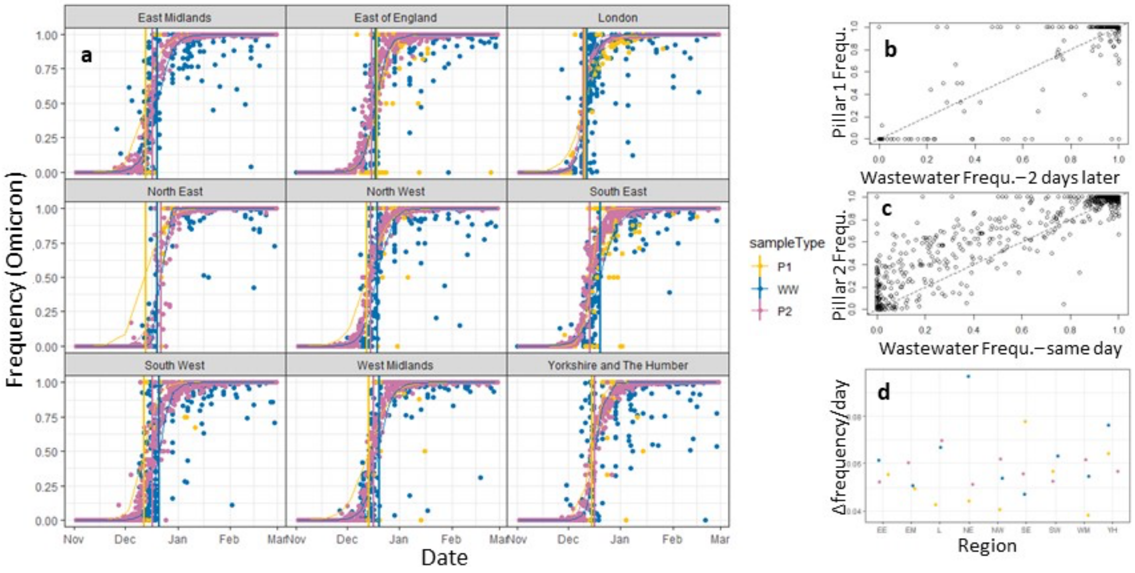


Figure S7: **a** Log-logistic growth models of the Omicron frequencies in 9 English regions. Vertical lines indicate the inflection points of the models (e), which estimate the timepoints when Omicron frequencies reach 50% in wastewater (blue), individual testing in clinical settings (Pillar 1, yellow) and individual testing in the wider population (Pillar 2, purple) samples. **b&c** Spearman's correlation tests show strongest correlation between county level daily Omicron frequency in individual testing and wastewater with a lag of 2 days in the wastewater compared to Pillar 1 (b) and with no lag compared to Pillar 2 (c, Table S1b). **d** growth rates of the log-logistic growth models in each region in wastewater (blue), Pillar 1 (yellow) and Pillar 2 (purple) samples, calculated as predicted frequency change between 5 days before and 5 days after inflection point.

Table S1a: Correlations between Omicron frequencies in combined individual testing and wastewater samples

| Wastewater timing relative to individual testing | Spearman's $\rho$ | n | P |
|---|---|---|---|
| Lag 5 days | 0.876 | 2010 | <0.001 |
| Lag 4 days | 0.886 | 2012 | <0.001 |
| Lag 3 days | 0.894 | 2055 | <0.001 |
| Lag 2 days | 0.897 | 2067 | <0.001 |
| Lag 1 day | 0.898 | 2083 | <0.001 |
| Same day | **0.905** | 2096 | <0.001 |
| Lead 1 day | 0.896 | 2097 | <0.001 |
| Lead 2 days | 0.897 | 2060 | <0.001 |
| Lead 3 days | 0.899 | 2045 | <0.001 |

Table S1b: Correlations between Omicron frequencies in pillar 1 or pillar 2 individual testing and wastewater samples

| Wastewater timing lead /lag | Pillar 1: Spearman's $\rho$ | Pillar 1: n | Pillar 1: P | Pillar 2: Spearman's $\rho$ | Pillar 2: n | Pillar 2: P |
|---|---|---|---|---|---|---|
| Lag 5 | 0.860 | 682 | <0.001 | 0.880 | 2010 | <0.001 |
| Lag 4 | 0.856 | 722 | <0.001 | 0.890 | 2012 | <0.001 |
| Lag 3 | 0.866 | 755 | <0.001 | 0.899 | 2055 | <0.001 |
| Lag 2 | **0.871** | 772 | <0.001 | 0.901 | 2067 | <0.001 |
| Lag 1 | 0.861 | 731 | <0.001 | 0.903 | 2083 | <0.001 |
| 0 | 0.857 | 710 | <0.001 | **0.908** | 2096 | <0.001 |

| | | | | | | |
|---|---|---|---|---|---|---|
| Lead 1 | 0.855 | 713 | <0.001 | 0.902 | 2097 | <0.001 |
| Lead 2 | 0.823 | 697 | <0.001 | 0.904 | 2060 | <0.001 |
| Lead 3 | 0.844 | 746 | <0.001 | 0.902 | 2045 | <0.001 |

### The COVID-19 Genomics UK (COG-UK) consortium

**June 2021 V.3**

**Funding acquisition, Leadership and supervision, Metadata curation, Project administration, Samples and logistics, Sequencing and analysis, Software and analysis tools, and Visualisation:**
Dr Samuel C Robson PhD [13, 84]

**Funding acquisition, Leadership and supervision, Metadata curation, Project administration, Samples and logistics, Sequencing and analysis, and Software and analysis tools:**
Dr Thomas R Connor PhD [11, 74] and Prof Nicholas J Loman PhD [43]

**Leadership and supervision, Metadata curation, Project administration, Samples and logistics, Sequencing and analysis, Software and analysis tools, and Visualisation:**
Dr Tanya Golubchik PhD [5]

**Funding acquisition, Leadership and supervision, Metadata curation, Samples and logistics, Sequencing and analysis, and Visualisation:**
Dr Rocio T Martinez Nunez PhD [46]

**Funding acquisition, Leadership and supervision, Project administration, Samples and logistics, Sequencing and analysis, and Software and analysis tools:**
Dr David Bonsall PhD [5]

**Funding acquisition, Leadership and supervision, Project administration, Sequencing and analysis, Software and analysis tools, and Visualisation:**
Prof Andrew Rambaut DPhil [104]

**Funding acquisition, Metadata curation, Project administration, Samples and logistics, Sequencing and analysis, and Software and analysis tools:**
Dr Luke B Snell MSc, MBBS [12]

**Leadership and supervision, Metadata curation, Project administration, Samples and logistics, Software and analysis tools, and Visualisation:**
Rich Livett MSc [116]

**Funding acquisition, Leadership and supervision, Metadata curation, Project administration, and Samples and logistics:**
Dr Catherine Ludden PhD [20, 70]

**Funding acquisition, Leadership and supervision, Metadata curation, Samples and logistics, and Sequencing and analysis:**

Dr Sally Corden PhD [74] and Dr Eleni Nastouli FRCPath [96, 95, 30]

**Funding acquisition, Leadership and supervision, Metadata curation, Sequencing and analysis, and Software and analysis tools:**
Dr Gaia Nebbia PhD, FRCPath [12]

**Funding acquisition, Leadership and supervision, Project administration, Samples and logistics, and Sequencing and analysis:**
Ian Johnston BSc [116]

**Leadership and supervision, Metadata curation, Project administration, Samples and logistics, and Sequencing and analysis:**
Prof Katrina Lythgoe PhD [5], Dr M. Estee Torok FRCP [19, 20] and Prof Ian G Goodfellow PhD [24]

**Leadership and supervision, Metadata curation, Project administration, Samples and logistics, and Visualisation:**
Dr Jacqui A Prieto PhD [97, 82] and Dr Kordo Saeed MD, FRCPath [97, 83]

**Leadership and supervision, Metadata curation, Project administration, Sequencing and analysis, and Software and analysis tools:**
Dr David K Jackson PhD [116]

**Leadership and supervision, Metadata curation, Samples and logistics, Sequencing and analysis, and Visualisation:**
Dr Catherine Houlihan PhD [96, 94]

**Leadership and supervision, Metadata curation, Sequencing and analysis, Software and analysis tools, and Visualisation:**
Dr Dan Frampton PhD [94, 95]

**Metadata curation, Project administration, Samples and logistics, Sequencing and analysis, and Software and analysis tools:**
Dr William L Hamilton PhD [19] and Dr Adam A Witney PhD [41]

**Funding acquisition, Samples and logistics, Sequencing and analysis, and Visualisation:**
Dr Giselda Bucca PhD [101]

**Funding acquisition, Leadership and supervision, Metadata curation, and Project administration:**
Dr Cassie F Pope PhD[40, 41]

**Funding acquisition, Leadership and supervision, Metadata curation, and Samples and logistics:**
Dr Catherine Moore PhD [74]

**Funding acquisition, Leadership and supervision, Metadata curation, and Sequencing and analysis:**
Prof Emma C Thomson PhD, FRCP [53]

**Funding acquisition, Leadership and supervision, Project administration, and Samples and logistics:**
Dr Teresa Cutino-Moguel PhD [2], Dr Ewan M Harrison PhD [116, 102]

**Funding acquisition, Leadership and supervision, Sequencing and analysis, and Visualisation:**
Prof Colin P Smith PhD [101]

**Leadership and supervision, Metadata curation, Project administration, and Sequencing and analysis:**
Fiona Rogan BSc [77]

**Leadership and supervision, Metadata curation, Project administration, and Samples and logistics:**
Shaun M Beckwith MSc [6], Abigail Murray Degree [6], Dawn Singleton HNC [6], Dr Kirstine Eastick PhD, FRCPath [37], Dr Liz A Sheridan PhD [98], Paul Randell MSc, PgD [99], Dr Leigh M Jackson PhD [105], Dr Cristina V Ariani PhD [116] and Dr Sónia Gonçalves PhD [116]

**Leadership and supervision, Metadata curation, Samples and logistics, and Sequencing and analysis:**
Dr Derek J Fairley PhD [3, 77], Prof Matthew W Loose PhD [18] and Joanne Watkins MSc [74]

**Leadership and supervision, Metadata curation, Samples and logistics, and Visualisation:**
Dr Samuel Moses MD [25, 106]

**Leadership and supervision, Metadata curation, Sequencing and analysis, and Software and analysis tools:**
Dr Sam Nicholls PhD [43], Dr Matthew Bull PhD [74] and Dr Roberto Amato PhD [116]

**Leadership and supervision, Project administration, Samples and logistics, and Sequencing and analysis:**
Prof Darren L Smith PhD [36, 65, 66]

**Leadership and supervision, Sequencing and analysis, Software and analysis tools, and Visualisation:**
Prof David M Aanensen PhD [14, 116] and Dr Jeffrey C Barrett PhD [116]

**Metadata curation, Project administration, Samples and logistics, and Sequencing and analysis:**
Dr Beatrix Kele PhD [2], Dr Dinesh Aggarwal MRCP[20, 116, 70], Dr James G Shepherd MBCHB, MRCP [53], Dr Martin D Curran PhD [71] and Dr Surendra Parmar PhD [71]

**Metadata curation, Project administration, Sequencing and analysis, and Software and analysis tools:**
Dr Matthew D Parker PhD [109]

**Metadata curation, Samples and logistics, Sequencing and analysis, and Software and analysis tools:**
Dr Catryn Williams PhD [74]

**Metadata curation, Samples and logistics, Sequencing and analysis, and Visualisation:**
Dr Sharon Glaysher PhD [68]

**Metadata curation, Sequencing and analysis, Software and analysis tools, and Visualisation:**
Dr Anthony P Underwood PhD [14, 116], Dr Matthew Bashton PhD [36, 65], Dr Nicole Pacchiarini PhD [74], Dr Katie F Loveson PhD [84] and Matthew Byott MSc [95, 96]

**Project administration, Sequencing and analysis, Software and analysis tools, and Visualisation:**
Dr Alessandro M Carabelli PhD [20]

**Funding acquisition, Leadership and supervision, and Metadata curation:**
Dr Kate E Templeton PhD [56, 104]

**Funding acquisition, Leadership and supervision, and Project administration:**
Dr Thushan I de Silva PhD [109], Dr Dennis Wang PhD [109], Dr Cordelia F Langford PhD [116] and John Sillitoe BEng [116]

**Funding acquisition, Leadership and supervision, and Samples and logistics:**
Prof Rory N Gunson PhD, FRCPath [55]

**Funding acquisition, Leadership and supervision, and Sequencing and analysis:**
Dr Simon Cottrell PhD [74], Dr Justin O'Grady PhD [75, 103] and Prof Dominic Kwiatkowski PhD [116, 108]

**Leadership and supervision, Metadata curation, and Project administration:**
Dr Patrick J Lillie PhD, FRCP [37]

**Leadership and supervision, Metadata curation, and Samples and logistics:**
Dr Nicholas Cortes MBCHB [33], Dr Nathan Moore MBCHB [33], Dr Claire Thomas DPhil [33], Phillipa J Burns MSc, DipRCPath [37], Dr Tabitha W Mahungu FRCPath [80] and Steven Liggett BSc [86]

**Leadership and supervision, Metadata curation, and Sequencing and analysis:**
Angela H Beckett MSc [13, 81] and Prof Matthew TG Holden PhD [73]

**Leadership and supervision, Project administration, and Samples and logistics:**
Dr Lisa J Levett PhD [34], Dr Husam Osman PhD [70, 35] and Dr Mohammed O Hassan-Ibrahim PhD, FRCPath [99]

**Leadership and supervision, Project administration, and Sequencing and analysis:**
Dr David A Simpson PhD [77]

**Leadership and supervision, Samples and logistics, and Sequencing and analysis:**
Dr Meera Chand PhD [72], Prof Ravi K Gupta PhD [102], Prof Alistair C Darby PhD [107] and Prof Steve Paterson PhD [107]

**Leadership and supervision, Sequencing and analysis, and Software and analysis tools:**
Prof Oliver G Pybus DPhil [23], Dr Erik M Volz PhD [39], Prof Daniela de Angelis PhD [52], Prof David L Robertson PhD [53], Dr Andrew J Page PhD [75] and Dr Inigo Martincorena PhD [116]

**Leadership and supervision, Sequencing and analysis, and Visualisation:**
Dr Louise Aigrain PhD [116] and Dr Andrew R Bassett PhD [116]

**Metadata curation, Project administration, and Samples and logistics:**
Dr Nick Wong DPhil, MRCP, FRCPath [50], Dr Yusri Taha MD, PhD [89], Michelle J Erkiert BA [99] and Dr Michael H Spencer Chapman MBBS [116, 102]

**Metadata curation, Project administration, and Sequencing and analysis:**
Dr Rebecca Dewar PhD [56] and Martin P McHugh MSc [56, 111]

**Metadata curation, Project administration, and Software and analysis tools:**
Siddharth Mookerjee MPH [38, 57]

**Metadata curation, Project administration, and Visualisation:**
Stephen Aplin [97], Matthew Harvey [97], Thea Sass [97], Dr Helen Umpleby FRCP [97] and Helen Wheeler [97]

**Metadata curation, Samples and logistics, and Sequencing and analysis:**
Dr James P McKenna PhD [3], Dr Ben Warne MRCP [9], Joshua F Taylor MSc [22], Yasmin Chaudhry BSc [24], Rhys Izuagbe [24], Dr Aminu S Jahun PhD [24], Dr Gregory R Young PhD [36, 65], Dr Claire McMurray PhD [43], Dr Clare M McCann PhD [65, 66], Dr Andrew Nelson PhD [65, 66] and Scott Elliott [68]

**Metadata curation, Samples and logistics, and Visualisation:**
Hannah Lowe MSc [25]

**Metadata curation, Sequencing and analysis, and Software and analysis tools:**
Dr Anna Price PhD [11], Matthew R Crown BSc [65], Dr Sara Rey PhD [74], Dr Sunando Roy PhD [96] and Dr Ben Temperton PhD [105]

**Metadata curation, Sequencing and analysis, and Visualisation:**
Dr Sharif Shaaban PhD [73] and Dr Andrew R Hesketh PhD [101]

**Project administration, Samples and logistics, and Sequencing and analysis:**
Dr Kenneth G Laing PhD[41], Dr Irene M Monahan PhD [41] and Dr Judith Heaney PhD [95, 96, 34]

**Project administration, Samples and logistics, and Visualisation:**
Dr Emanuela Pelosi FRCPath [97], Siona Silviera MSc [97] and Dr Eleri Wilson-Davies MD, FRCPath [97]

**Samples and logistics, Software and analysis tools, and Visualisation:**
Dr Helen Fryer PhD [5]

**Sequencing and analysis, Software and analysis tools, and Visualization:**
Dr Helen Adams PhD [4], Dr Louis du Plessis PhD [23], Dr Rob Johnson PhD [39], Dr William T Harvey PhD [53, 42], Dr Joseph Hughes PhD [53], Dr Richard J Orton PhD [53], Dr Lewis G Spurgin PhD [59], Dr Yann Bourgeois PhD [81], Dr Chris Ruis PhD [102], Áine O'Toole MSc [104], Marina Gourtovaia MSc [116] and Dr Theo Sanderson PhD [116]

**Funding acquisition, and Leadership and supervision:**
Dr Christophe Fraser PhD [5], Dr Jonathan Edgeworth PhD, FRCPath [12], Prof Judith Breuer MD [96, 29], Dr Stephen L Michell PhD [105] and Prof John A Todd PhD [115]

**Funding acquisition, and Project administration:**
Michaela John BSc [10] and Dr David Buck PhD [115]

**Leadership and supervision, and Metadata curation:**
Dr Kavitha Gajee MBBS, FRCPath [37] and Dr Gemma L Kay PhD [75]

**Leadership and supervision, and Project administration:**
Prof Sharon J Peacock PhD [20, 70] and David Heyburn [74]

**Leadership and supervision, and Samples and logistics:**
Dr Themoula Charalampous PhD [12, 46], Adela Alcolea-Medina [32, 112], Katie Kitchman BSc [37], Prof Alan McNally PhD [43, 93], David T Pritchard MSc, CSci [50], Dr Samir Dervisevic FRCPath [58], Dr Peter Muir PhD [70], Dr Esther Robinson PhD [70, 35], Dr Barry B Vipond PhD [70], Newara A Ramadan MSc, CSci, FIBMS [78], Dr Christopher Jeanes MBBS [90], Danni Weldon BSc [116], Jana Catalan MSc [118] and Neil Jones MSc [118]

**Leadership and supervision, and Sequencing and analysis:**
Dr Ana da Silva Filipe PhD [53], Dr Chris Williams MBBS [74], Marc Fuchs BSc [77], Dr Julia Miskelly PhD [77], Dr Aaron R Jeffries PhD [105], Karen Oliver BSc [116] and Dr Naomi R Park PhD [116]

**Metadata curation, and Samples and logistics:**
Amy Ash BSc [1], Cherian Koshy MSc, CSci, FIBMS [1], Magdalena Barrow [7], Dr Sarah L Buchan PhD [7], Dr Anna Mantzouratou PhD [7], Dr Gemma Clark PhD [15], Dr Christopher W Holmes PhD [16], Sharon Campbell MSc [17], Thomas Davis MSc [21], Ngee Keong Tan MSc [22], Dr Julianne R Brown PhD [29], Dr Kathryn A Harris PhD [29, 2], Stephen P Kidd MSc [33], Dr Paul R Grant PhD [34], Dr Li Xu-McCrae PhD [35], Dr Alison Cox PhD [38, 63], Pinglawathee Madona [38, 63], Dr Marcus Pond PhD [38, 63], Dr Paul A Randell MBBCh [38, 63], Karen T Withell FIBMS [48], Cheryl Williams MSc [51], Dr Clive Graham MD [60], Rebecca Denton-Smith BSc [62], Emma Swindells BSc [62], Robyn Turnbull BSc [62], Dr Tim J Sloan PhD [67], Dr Andrew Bosworth PhD [70, 35], Stephanie Hutchings [70], Hannah M Pymont MSc [70], Dr Anna Casey PhD [76], Dr Liz Ratcliffe PhD [76], Dr Christopher R Jones PhD [79, 105], Dr Bridget A Knight PhD [79, 105], Dr Tanzina Haque PhD, FRCPath [80], Dr Jennifer Hart MRCP [80], Dr Dianne Irish-Tavares FRCPath [80], Eric Witele MSc [80], Craig Mower BA [86], Louisa K Watson DipHE [86], Jennifer Collins BSc [89], Gary Eltringham BSc [89], Dorian Crudgington [98], Ben Macklin [98], Prof Miren Iturriza-Gomara PhD [107], Dr Anita O Lucaci PhD [107] and Dr Patrick C McClure PhD [113]

**Metadata curation, and Sequencing and analysis:**
Matthew Carlile BSc [18], Dr Nadine Holmes PhD [18], Dr Christopher Moore PhD [18], Dr Nathaniel Storey PhD [29], Dr Stefan Rooke PhD [73], Dr Gonzalo Yebra PhD [73], Dr Noel Craine DPhil [74], Malorie Perry MSc [74], Dr Nabil-Fareed Alikhan PhD [75], Dr Stephen Bridgett PhD [77], Kate F Cook MScR [84], Christopher Fearn MSc [84], Dr Salman Goudarzi PhD [84], Prof Ronan A Lyons MD [88], Dr Thomas Williams MD [104], Dr Sam T Haldenby PhD [107], Jillian Durham BSc [116] and Dr Steven Leonard PhD [116]

**Metadata curation, and Software and analysis tools:**
Robert M Davies MA (Cantab) [116]

**Project administration, and Samples and logistics:**
Dr Rahul Batra MD [12], Beth Blane BSc [20], Dr Moira J Spyer PhD [30, 95, 96], Perminder Smith MSc [32, 112], Mehmet Yavus [85, 109], Dr Rachel J Williams PhD [96], Dr Adhyana IK Mahanama MD [97], Dr Buddhini Samaraweera MD [97], Sophia T Girgis MSc [102], Samantha E Hansford CSci [109], Dr Angie Green PhD [115], Dr Charlotte Beaver PhD [116], Katherine L Bellis [116, 102], Matthew J Dorman [116], Sally Kay [116], Liam Prestwood [116] and Dr Shavanthi Rajatileka PhD [116]

**Project administration, and Sequencing and analysis:**
Dr Joshua Quick PhD [43]

**Project administration, and Software and analysis tools:**
Radoslaw Poplawski BSc [43]

**Samples and logistics, and Sequencing and analysis:**
Dr Nicola Reynolds PhD [8], Andrew Mack MPhil [11], Dr Arthur Morriss PhD [11], Thomas Whalley BSc [11], Bindi Patel BSc [12], Dr Iliana Georgana PhD [24], Dr Myra Hosmillo PhD [24], Malte L Pinckert MPhil [24], Dr Joanne Stockton PhD [43], Dr John H Henderson PhD [65], Amy Hollis HND [65], Dr William Stanley PhD [65], Dr Wen C Yew PhD [65], Dr Richard Myers PhD [72], Dr Alicia Thornton PhD [72], Alexander Adams BSc [74], Tara Annett BSc [74], Dr Hibo Asad PhD [74], Alec Birchley MSc [74], Jason Coombes BSc [74], Johnathan M Evans MSc [74], Laia Fina [74], Bree Gatica-Wilcox MPhil [74], Lauren Gilbert [74], Lee Graham BSc [74], Jessica Hey BSc [74], Ember Hilvers MPH [74], Sophie Jones MSc [74], Hannah Jones [74], Sara Kumziene-

Summerhayes MSc [74], Dr Caoimhe McKerr PhD [74], Jessica Powell BSc [74], Georgia Pugh [74], Sarah Taylor [74], Alexander J Trotter MRes [75], Charlotte A Williams BSc [96], Leanne M Kermack MSc [102], Benjamin H Foulkes MSc [109], Marta Gallis MSc [109], Hailey R Hornsby MSc [109], Stavroula F Louka MSc [109], Dr Manoj Pohare PhD [109], Paige Wolverson MSc [109], Peijun Zhang MSc [109], George MacIntyre-Cockett BSc [115], Amy Trebes MSc [115], Dr Robin J Moll PhD [116], Lynne Ferguson MSc [117], Dr Emily J Goldstein PhD [117], Dr Alasdair Maclean PhD [117] and Dr Rachael Tomb PhD [117]

**Samples and logistics, and Software and analysis tools:**
Dr Igor Starinskij MSc, MRCP [53]

**Sequencing and analysis, and Software and analysis tools:**
Laura Thomson BSc [5], Joel Southgate MSc [11, 74], Dr Moritz UG Kraemer DPhil [23], Dr Jayna Raghwani PhD [23], Dr Alex E Zarebski PhD [23], Olivia Boyd MSc [39], Lily Geidelberg MSc [39], Dr Chris J Illingworth PhD [52], Dr Chris Jackson PhD [52], Dr David Pascall PhD [52], Dr Sreenu Vattipally PhD [53], Timothy M Freeman MPhil [109], Dr Sharon N Hsu PhD [109], Dr Benjamin B Lindsey MRCP [109], Dr Keith James PhD [116], Kevin Lewis [116], Gerry Tonkin-Hill [116] and Dr Jaime M Tovar-Corona PhD [116]

**Sequencing and analysis, and Visualisation:**
MacGregor Cox MSci [20]

**Software and analysis tools, and Visualisation:**
Dr Khalil Abudahab PhD [14, 116], Mirko Menegazzo [14], Ben EW Taylor MEng [14, 116], Dr Corin A Yeats PhD [14], Afrida Mukaddas BTech [53], Derek W Wright MSc [53], Dr Leonardo de Oliveira Martins PhD [75], Dr Rachel Colquhoun DPhil [104], Verity Hill [104], Dr Ben Jackson PhD [104], Dr JT McCrone PhD [104], Dr Nathan Medd PhD [104], Dr Emily Scher PhD [104] and Jon-Paul Keatley [116]

**Leadership and supervision:**
Dr Tanya Curran PhD [3], Dr Sian Morgan FRCPath [10], Prof Patrick Maxwell PhD [20], Prof Ken Smith PhD [20], Dr Sahar Eldirdiri MBBS, MSc, FRCPath [21], Anita Kenyon MSc [21], Prof Alison H Holmes MD [38, 57], Dr James R Price PhD [38, 57], Dr Tim Wyatt PhD [69], Dr Alison E Mather PhD [75], Dr Timofey Skvortsov PhD [77] and Prof John A Hartley PhD [96]

**Metadata curation:**
Prof Martyn Guest PhD [11], Dr Christine Kitchen PhD [11], Dr Ian Merrick PhD [11], Robert Munn BSc [11], Dr Beatrice Bertolusso Degree [33], Dr Jessica Lynch MBCHB [33], Dr Gabrielle Vernet MBBS [33], Stuart Kirk MSc [34], Dr Elizabeth Wastnedge MD [56], Dr Rachael Stanley PhD [58], Giles Idle [64], Dr Declan T Bradley PhD [69, 77], Nicholas F Killough MSc [69], Dr Jennifer Poyner MD [79] and Matilde Mori BSc [110]

**Project administration:**
Owen Jones BSc [11], Victoria Wright BSc [18], Ellena Brooks MA [20], Carol M Churcher BSc [20], Mireille Fragakis HND [20], Dr Katerina Galai PhD [20, 70], Dr Andrew Jermy PhD [20], Sarah Judges BA [20], Georgina M McManus BSc [20], Kim S Smith [20], Dr Elaine Westwick PhD [20], Dr Stephen W Attwood PhD [23], Dr Frances Bolt PhD [38, 57], Dr Alisha Davies PhD [74], Elen De Lacy MPH [74], Fatima Downing [74], Sue Edwards [74], Lizzie Meadows MA [75], Sarah Jeremiah MSc [97], Dr Nikki Smith PhD [109] and Luke Foulser [116]

**Samples and logistics:**
Amita Patel BSc [12], Dr Louise Berry PhD [15], Dr Tim Boswell PhD [15], Dr Vicki M Fleming PhD [15], Dr Hannah C Howson-Wells PhD [15], Dr Amelia Joseph PhD [15], Manjinder Khakh [15], Dr Michelle M Lister PhD [15], Paul W Bird MSc, MRes [16], Karlie Fallon [16], Thomas Helmer [16], Dr Claire L McMurray PhD [16], Mina Odedra BSc [16], Jessica Shaw BSc [16], Dr Julian W Tang PhD [16], Nicholas J Willford MSc [16], Victoria Blakey BSc [17], Dr Veena Raviprakash MD [17], Nicola Sheriff BSc [17], Lesley-Anne Williams BSc [17], Theresa

Feltwell MSc [20], Dr Luke Bedford PhD [26], Dr James S Cargill PhD [27], Warwick Hughes MSc [27], Dr Jonathan Moore MD [28], Susanne Stonehouse BSc [28], Laura Atkinson MSc [29], Jack CD Lee MSc [29], Dr Divya Shah PhD [29], Natasha Ohemeng-Kumi MSc [32, 112], John Ramble MSc [32, 112], Jasveen Sehmi MSc [32, 112], Dr Rebecca Williams BMBS [33], Wendy Chatterton MSc [34], Monika Pusok MSc [34], William Everson MSc [37], Anibolina Castigador IBMS HCPC [44], Emily Macnaughton FRCPath [44], Dr Kate El Bouzidi MRCP [45], Dr Temi Lampejo FRCPath [45], Dr Malur Sudhanva FRCPath [45], Cassie Breen BSc [47], Dr Graciela Sluga MD, MSc [48], Dr Shazaad SY Ahmad MSc [49, 70], Dr Ryan P George PhD [49], Dr Nicholas W Machin MSc [49, 70], Debbie Binns BSc [50], Victoria James BSc [50], Dr Rachel Blacow MBCHB [55], Dr Lindsay Coupland PhD [58], Dr Louise Smith PhD [59], Dr Edward Barton MD [60], Debra Padgett BSc [60], Garren Scott BSc [60], Dr Aidan Cross MBCHB [61], Dr Mariyam Mirfenderesky FRCPath [61], Jane Greenaway MSc [62], Kevin Cole [64], Phillip Clarke [67], Nichola Duckworth [67], Sarah Walsh [67], Kelly Bicknell [68], Robert Impey MSc [68], Dr Sarah Wyllie PhD [68], Richard Hopes [70], Dr Chloe Bishop PhD [72], Dr Vicki Chalker PhD [72], Dr Ian Harrison PhD [72], Laura Gifford MSc [74], Dr Zoltan Molnar PhD [77], Dr Cressida Auckland FRCPath [79], Dr Cariad Evans PhD [85, 109], Dr Kate Johnson PhD [85, 109], Dr David G Partridge FRCP, FRCPath [85, 109], Dr Mohammad Raza PhD [85, 109], Paul Baker MD [86], Prof Stephen Bonner PhD [86], Sarah Essex [86], Leanne J Murray [86], Andrew I Lawton MSc [87], Dr Shirelle Burton-Fanning MD [89], Dr Brendan AI Payne MD [89], Dr Sheila Waugh MD [89], Andrea N Gomes MSc [91], Maimuna Kimuli MSc [91], Darren R Murray MSc [91], Paula Ashfield MSc [92], Dr Donald Dobie MBCHB [92], Dr Fiona Ashford PhD [93], Dr Angus Best PhD [93], Dr Liam Crawford PhD [93], Dr Nicola Cumley PhD [93], Dr Megan Mayhew PhD [93], Dr Oliver Megram PhD [93], Dr Jeremy Mirza PhD [93], Dr Emma Moles-Garcia PhD [93], Dr Benita Percival PhD [93], Megan Driscoll BSc [96], Leah Ensell BSc [96], Dr Helen L Lowe PhD [96], Laurentiu Maftei BSc [96], Matteo Mondani MSc [96], Nicola J Chaloner BSc [99], Benjamin J Cogger BSc [99], Lisa J Easton MSc [99], Hannah Huckson BSc [99], Jonathan Lewis MSc, PgD, FIBMS [99], Sarah Lowdon BSc [99], Cassandra S Malone MSc [99], Florence Munemo BSc [99], Manasa Mutingwende MSc [99], Roberto Nicodemi BSc [99], Olga Podplomyk FD [99], Thomas Somassa BSc [99], Dr Andrew Beggs PhD [100], Dr Alex Richter PhD [100], Claire Cormie [102], Joana Dias MSc [102], Sally Forrest BSc [102], Dr Ellen E Higginson PhD [102], Mailis Maes MPhil [102], Jamie Young BSc [102], Dr Rose K Davidson PhD [103], Kathryn A Jackson MSc [107], Dr Alexander J Keeley MRCP [109], Prof Jonathan Ball PhD [113], Timothy Byaruhanga MSc [113], Dr Joseph G Chappell PhD [113], Jayasree Dey MSc [113], Jack D Hill MSc [113], Emily J Park MSc [113], Arezou Fanaie MSc [114], Rachel A Hilson MSc [114], Geraldine Yaze MSc [114] and Stephanie Lo [116]

**Sequencing and analysis:**

Safiah Afifi BSc [10], Robert Beer BSc [10], Joshua Maksimovic FD [10], Kathryn McCluggage Masters [10], Karla Spellman FD [10], Catherine Bresner BSc [11], William Fuller BSc [11], Dr Angela Marchbank BSc [11], Trudy Workman HNC [11], Dr Ekaterina Shelest PhD [13, 81], Dr Johnny Debebe PhD [18], Dr Fei Sang PhD [18], Dr Sarah Francois PhD [23], Bernardo Gutierrez MSc [23], Dr Tetyana I Vasylyeva DPhil [23], Dr Flavia Flaviani PhD [31], Dr Manon Ragonnet-Cronin PhD [39], Dr Katherine L Smollett PhD [42], Alice Broos BSc [53], Daniel Mair BSc [53], Jenna Nichols BSc [53], Dr Kyriaki Nomikou PhD [53], Dr Lily Tong PhD [53], Ioulia Tsatsani MSc [53], Prof Sarah O'Brien PhD [54], Prof Steven Rushton PhD [54], Dr Roy Sanderson PhD [54], Dr Jon Perkins MBCHB [55], Seb Cotton MSc [56], Abbie Gallagher BSc [56], Dr Elias Allara MD, PhD [70, 102], Clare Pearson MSc [70, 102], Dr David Bibby PhD [72], Dr Gavin Dabrera PhD [72], Dr Nicholas Ellaby PhD [72], Dr Eileen Gallagher PhD [72], Dr Jonathan Hubb PhD [72], Dr Angie Lackenby PhD [72], Dr David Lee PhD [72], Nikos Manesis [72], Dr Tamyo Mbisa PhD [72], Dr Steven Platt PhD [72], Katherine A Twohig [72], Dr Mari Morgan PhD [74], Alp Aydin MSci [75], David J Baker BEng [75], Dr Ebenezer Foster-Nyarko PhD [75], Dr Sophie J Prosolek PhD [75], Steven Rudder [75], Chris Baxter BSc [77], Sílvia F Carvalho MSc [77], Dr Deborah Lavin PhD [77], Dr Arun Mariappan PhD [77], Dr Clara Radulescu PhD [77], Dr Aditi Singh PhD [77], Miao Tang MD [77], Helen Morcrette BSc [79], Nadua Bayzid BSc [96], Marius Cotic MSc [96], Dr Carlos E Balcazar PhD [104], Dr Michael D Gallagher PhD [104], Dr Daniel Maloney PhD [104], Thomas D Stanton BSc [104], Dr Kathleen A Williamson PhD [104], Dr Robin Manley PhD [105], Michelle L Michelsen BSc [105], Dr Christine M Sambles PhD [105], Dr David J Studholme PhD [105], Joanna Warwick-Dugdale BSc [105], Richard Eccles MSc [107], Matthew Gemmell MSc [107], Dr Richard Gregory PhD [107], Dr Margaret Hughes PhD [107], Charlotte

Nelson MSc [107], Dr Lucille Rainbow PhD [107], Dr Edith E Vamos PhD [107], Hermione J Webster BSc [107], Dr Mark Whitehead PhD [107], Claudia Wierzbicki BSc [107], Dr Adrienn Angyal PhD [109], Dr Luke R Green PhD [109], Dr Max Whiteley PhD [109], Emma Betteridge BSc [116], Dr Iraad F Bronner PhD [116], Ben W Farr BSc [116], Scott Goodwin MSc [116], Dr Stefanie V Lensing PhD [116], Shane A McCarthy [116, 102], Dr Michael A Quail PhD [116], Diana Rajan MSc [116], Dr Nicholas M Redshaw PhD [116], Carol Scott [116], Lesley Shirley MSc [116] and Scott AJ Thurston BSc [116]

**Software and analysis tools:**
Dr Will Rowe PhD[43], Amy Gaskin MSc [74], Dr Thanh Le-Viet PhD [75], James Bonfield BSc [116], Jennifier Liddle [116] and Andrew Whitwham BSc [116]

**1** Barking, Havering and Redbridge University Hospitals NHS Trust, **2** Barts Health NHS Trust, **3** Belfast Health & Social Care Trust, **4** Betsi Cadwaladr University Health Board, **5** Big Data Institute, Nuffield Department of Medicine, University of Oxford, **6** Blackpool Teaching Hospitals NHS Foundation Trust, **7** Bournemouth University, **8** Cambridge Stem Cell Institute, University of Cambridge, **9** Cambridge University Hospitals NHS Foundation Trust, **10** Cardiff and Vale University Health Board, **11** Cardiff University, **12** Centre for Clinical Infection and Diagnostics Research, Department of Infectious Diseases, Guy's and St Thomas' NHS Foundation Trust, **13** Centre for Enzyme Innovation, University of Portsmouth, **14** Centre for Genomic Pathogen Surveillance, University of Oxford, **15** Clinical Microbiology Department, Queens Medical Centre, Nottingham University Hospitals NHS Trust, **16** Clinical Microbiology, University Hospitals of Leicester NHS Trust, **17** County Durham and Darlington NHS Foundation Trust, **18** Deep Seq, School of Life Sciences, Queens Medical Centre, University of Nottingham, **19** Department of Infectious Diseases and Microbiology, Cambridge University Hospitals NHS Foundation Trust, **20** Department of Medicine, University of Cambridge, **21** Department of Microbiology, Kettering General Hospital, **22** Department of Microbiology, South West London Pathology, **23** Department of Zoology, University of Oxford, **24** Division of Virology, Department of Pathology, University of Cambridge, **25** East Kent Hospitals University NHS Foundation Trust, **26** East Suffolk and North Essex NHS Foundation Trust, **27** East Sussex Healthcare NHS Trust**, 28** Gateshead Health NHS Foundation Trust, **29** Great Ormond Street Hospital for Children NHS Foundation Trust, **30** Great Ormond Street Institute of Child Health (GOS ICH), University College London (UCL), **31** Guy's and St. Thomas' Biomedical Research Centre, **32** Guy's and St. Thomas' NHS Foundation Trust, **33** Hampshire Hospitals NHS Foundation Trust, **34** Health Services Laboratories, **35** Heartlands Hospital, Birmingham, **36** Hub for Biotechnology in the Built Environment, Northumbria University, **37** Hull University Teaching Hospitals NHS Trust, **38** Imperial College Healthcare NHS Trust, **39** Imperial College London, **40** Infection Care Group, St George's University Hospitals NHS Foundation Trust, **41** Institute for Infection and Immunity, St George's University of London, **42** Institute of Biodiversity, Animal Health & Comparative Medicine, **43** Institute of Microbiology and Infection, University of Birmingham, **44** Isle of Wight NHS Trust, **45** King's College Hospital NHS Foundation Trust, **46** King's College London, **47** Liverpool Clinical Laboratories, **48** Maidstone and Tunbridge Wells NHS Trust, **49** Manchester University NHS Foundation Trust, **50** Microbiology Department, Buckinghamshire Healthcare NHS Trust, **51** Microbiology, Royal Oldham Hospital, **52** MRC Biostatistics Unit, University of Cambridge, **53** MRC-University of Glasgow Centre for Virus Research, **54** Newcastle University, **55** NHS Greater Glasgow and Clyde, **56** NHS Lothian, **57** NIHR Health Protection Research Unit in HCAI and AMR, Imperial College London, **58** Norfolk and Norwich University Hospitals NHS Foundation Trust, **59** Norfolk County Council, **60** North Cumbria Integrated Care NHS Foundation Trust, **61** North Middlesex University Hospital NHS Trust, **62** North Tees and Hartlepool NHS Foundation Trust, **63** North West London Pathology, **64** Northumbria Healthcare NHS Foundation Trust, **65** Northumbria University, **66** NU-OMICS, Northumbria University, **67** Path Links, Northern Lincolnshire and Goole NHS Foundation Trust, **68** Portsmouth Hospitals University NHS Trust, **69** Public Health Agency, Northern Ireland, **70** Public Health England, **71** Public Health England, Cambridge, **72** Public Health England, Colindale, **73** Public Health Scotland, **74** Public Health Wales, **75** Quadram Institute Bioscience, **76** Queen Elizabeth Hospital, Birmingham, **77** Queen's University Belfast, **78** Royal Brompton and Harefield Hospitals, **79** Royal Devon and Exeter NHS Foundation Trust, **80** Royal Free London NHS Foundation Trust, **81** School of Biological Sciences, University of Portsmouth, **82** School of Health Sciences, University of Southampton, **83** School of Medicine, University of Southampton, **84** School of Pharmacy & Biomedical Sciences, University of Portsmouth, **85** Sheffield Teaching Hospitals NHS Foundation Trust, **86** South Tees Hospitals NHS Foundation Trust, **87** Southwest Pathology Services, **88** Swansea University, **89** The Newcastle upon Tyne Hospitals NHS Foundation Trust, **90** The Queen Elizabeth Hospital King's Lynn NHS Foundation Trust, **91** The Royal Marsden NHS Foundation Trust, **92** The Royal Wolverhampton NHS Trust, **93** Turnkey Laboratory, University of

Birmingham, **94** University College London Division of Infection and Immunity**, 95** University College London Hospital Advanced Pathogen Diagnostics Unit**, 96** University College London Hospitals NHS Foundation Trust, **97** University Hospital Southampton NHS Foundation Trust, **98** University Hospitals Dorset NHS Foundation Trust, **99** University Hospitals Sussex NHS Foundation Trust, **100** University of Birmingham, **101** University of Brighton, **102** University of Cambridge, **103** University of East Anglia, **104** University of Edinburgh, **105** University of Exeter, **106** University of Kent, **107** University of Liverpool, **108** University of Oxford, **109** University of Sheffield, **110** University of Southampton, **111** University of St Andrews, **112** Viapath, Guy's and St Thomas' NHS Foundation Trust, and King's College Hospital NHS Foundation Trust, **113** Virology, School of Life Sciences, Queens Medical Centre, University of Nottingham, **114** Watford General Hospital, **115** Wellcome Centre for Human Genetics, Nuffield Department of Medicine, University of Oxford, **116** Wellcome Sanger Institute, **117** West of Scotland Specialist Virology Centre, NHS Greater Glasgow and Clyde, **118** Whittington Health NHS Trust
